# Monitoring populations at increased risk for SARS-CoV-2 infection in the community

**DOI:** 10.1101/2021.09.02.21263017

**Authors:** Emma Pritchard, Joel Jones, Karina Vihta, Nicole Stoesser, Philippa C. Matthews, David W. Eyre, Thomas House, John I Bell, John N Newton, Jeremy Farrar, Derrick Crook, Susan Hopkins, Duncan Cook, Emma Rourke, Ruth Studley, Ian Diamond, Tim Peto, Koen B. Pouwels, A. Sarah Walker, the COVID-19 Infection Survey Team

## Abstract

**Background:** The COVID-19 pandemic is rapidly evolving, with emerging variants and fluctuating control policies. Real-time population screening and identification of groups in whom positivity is highest could help monitor spread and inform public health messaging and strategy.

**Methods:** To develop a real-time screening process, we included results from nose and throat swabs and questionnaires taken 19 July 2020-17 July 2021 in the UK’s national COVID-19 Infection Survey. Fortnightly, associations between SARS-CoV-2 positivity and 60 demographic and behavioural characteristics were estimated using logistic regression models adjusted for potential confounders, considering multiple testing, collinearity, and reverse causality.

**Findings:** Of 4,091,537 RT-PCR results from 482,677 individuals, 29,903 (0·73%) were positive. As positivity rose September-November 2020, rates were independently higher in younger ages, and those living in Northern England, major urban conurbations, more deprived areas, and larger households. Rates were also higher in those returning from abroad, and working in healthcare or outside of home. When positivity peaked December 2020-January 2021 (Alpha), high positivity shifted to southern geographical regions. With national vaccine roll-out from December 2020, positivity reduced in vaccinated individuals. Associations attenuated as rates decreased between February-May 2021. Rising positivity rates in June-July 2021 (Delta) were independently higher in younger, male, and unvaccinated groups. Few factors were consistently associated with positivity. 25/45 (56%) confirmed associations would have been detected later using 28-day rather than 14-day periods.

**Interpretation:** Population-level demographic and behavioural surveillance can be a valuable tool in identifying the varying characteristics driving current SARS-CoV-2 positivity, allowing monitoring to inform public health policy.

**Funding:** Department of Health and Social Care (UK), Welsh Government, Department of Health (on behalf of the Northern Ireland Government), Scottish Government, National Institute for Health Research.

## Introduction

To 31^st^ August 2021, there have been over 216·3 million SARS-CoV-2 cases worldwide.^1^ Disparities in COVID-19 risk and outcomes based on demographics and behaviours have been described in the UK^2, 3^ and globally,^4, 5^ but emerging variants^6^ coupled with varying control policies, including differential vaccine roll-out programmes, reinforce the need to monitor characteristics of individuals “at increased risk” for SARS-CoV-2 infection continuously. For example, identifying groups in whom newly identified variants of concern are spreading in the community may be vital in preventing widespread transmission. In England, since 26^th^ March 2020, there have been three national lockdowns, a tiered system^7^ with varying restrictions in smaller geographical areas, and various other restrictions between these,^8^ all affecting behaviour and risk of acquiring and spreading SARS-CoV-2. Finding societal factors or specific behaviours where these restrictions are less effective may aid policy development. With restrictions being relaxed in many countries, rapidly identifying groups where positivity is rising in real-time can help monitor spread and target advice.

High-quality surveillance is challenging, particularly given the large proportion of asymptomatic SARS-CoV-2-infected individuals,^9^ with a balance between missing important but potentially imprecisely estimated signals (false-negatives) and noise (false-positives). With large datasets containing many potential risk factors, multiple testing is inevitably problematic,^10^ but standard approaches to building regression models restricting to smaller numbers of hypothesised associated factors risks missing true signals with a rapidly evolving pathogen and societal responses. The cumulative effect of missing data across many risk factors can mean substantial proportions of the original sample are excluded from penalised regression or backwards elimination, losing power,^11^ and risking bias if missingness depends on outcome.^12^ A method allowing numerous variable parametrisations of many individual variables would therefore be useful, provided collinearity and confounding can be avoided.^13^

Using the Office for National Statistics (ONS) COVID-19 Infection Survey, a large community-based surveillance study, we therefore developed a process to monitor groups with highest SARS-CoV-2 positivity week by week.

## Methods

### Study design

The ONS COVID-19 Infection Survey is a large household survey with longitudinal follow-up (ISRCTN21086382; https://www.ndm.ox.ac.uk/covid-19/covid-19-infection-survey/protocol-and-information-sheets). Private households are randomly selected on a continuous basis from address lists and previous surveys to provide a representative sample across the UK. Following verbal consent, a study worker visited each household to take written informed consent for individuals aged ≥2 years (from parents/carers for those 2–15 years; those 10–15 years also provided written assent). The study received ethical approval from the South Central Berkshire B Research Ethics Committee (20/SC/0195).

Participants were asked about demographics, behaviours, work, and vaccination uptake (https://www.ndm.ox.ac.uk/covid-19/covid-19-infection-survey/case-record-forms). At the first visit, participants were asked for consent for optional follow-up visits every week for the next month, then monthly thereafter. At each visit, participants provided a nose and throat self-swab.

### Inclusion/exclusion criteria

This analysis included visits from 19^th^ July 2020-17^th^ July 2021 with a positive or negative swab result, including one visit per participant within each discrete fortnight in this period, namely the first test-positive visit, otherwise the last (negative) visit. This mimics repeated point-prevalence surveys, similar to the English Real-time Assessment of Community Transmission (REACT) study.^14^

### Outcome and exposures

The outcome was any SARS-CoV-2 PCR-positive swab in each fortnight. For exposures, we identified eight non-missing key potential confounders (“core” variables): sex, ethnicity (white vs non-white as relatively small numbers in the latter), age (years), geographical region (12 levels; 9 English regions and 3 devolved administrations: Wales, Scotland, Northern Ireland), rural/urban classification (major urban area, urban town/city, rural town, and rural village), deprivation percentile (derived separately for each country^15–18^), household size, and whether the household was multigenerational (details in **Supplementary Methods**).

We next defined 60 non-core “screening” variables that could dynamically identify those at increased risk of testing positive (**Supplementary Table 1**), from questions detailing participant’s current work/school status, including ability to social distance and patient-facing healthcare/social-care roles, current health status including COVID-19 vaccination and smoking, household and living environment, and contacts including with care homes, hospitals, and confirmed COVID-19 cases.

Although participants are tested predominantly monthly, most behavioural questions relate to the last 7 days. As some participants already know/think they have COVID-19 (from symptoms or testing outside the study) this could affect behaviours reported immediately before study tests, leading to reverse causality. The screening variables were therefore grouped into those most plausibly preceding any current infection (47 variables), or potentially modified through knowledge of recent prior infection (13 variables, including social/physical contacts, frequency of shopping and/or socialising, time spent in others homes/other people spent in participants’ homes; **Supplementary Table 1B**). For the latter, rather than the self-report at the included visit, we considered the maximum reported value across all visits in the preceding 35 days, excluding the included visit, and included only participants with at least one negative visit in the preceding 10-35 days.

### Statistical analysis

Within each fortnight, associations with the eight “core” characteristics were estimated using logistic regression (numbers included per fortnight in **Supplementary Table 2**). These characteristics were included in all subsequent models regardless of statistical significance. For geographic region, South West England was the reference as this had the lowest SARS-CoV-2 positivity across the study, facilitating identification of where infections were increasing. Given the large number of effect estimates over the 52-week study period (e.g. shown for urban/rural classification in **Supplementary Figure 1**), we summarised the importance of each characteristic over time using two properties simultaneously: 1) global (Wald) p-value and 2) overall effect size, the standard error-weighted mean effect estimate setting the reference to the level with lowest positivity in each fortnight^19^:

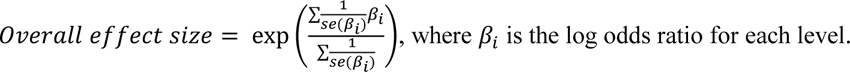

To incorporate non-linear effects, a restricted natural cubic spline was used for age (details in **Supplementary Methods**); the overall effect size combined estimates at ages 10, 25, 40, 55 vs 70 years (reference category) as above.

We tested interactions between the eight core variables individually in fortnights where positivity was >0·5% (arbitrary threshold to avoid small numbers), conducting backwards elimination on all with individual global heterogeneity p-value<0·001 (Bonferroni adjustment, 0·05/26 (number of interaction tests)), creating the “core model” (details in **Supplementary Methods**). An overall effect size was calculated for interactions as above, but taking the absolute coefficient values.

Given missing data (**Supplementary Table 1**), we used forward selection to retain as many participants as possible when screening each non-core characteristic, first adding each of the 47 “screening” variables individually to the “core model”, thus estimating the total effects not explained by core characteristics. For all work-related variables, work status was included regardless of significance so that effects reflected additional effects of the characteristic for those currently employed and working. To monitor multiple testing, we plotted observed p-values (global per variable and individual level vs reference) against expected p-values assuming no difference (randomly distributed between 0 and 1 given the number of tests), creating a Q-Q plot, including 0·05, Bonferroni and Benjamini-Hochberg adjusted p-values (0·05/tests) as references. As the goal was to identify signals of “at-risk” populations, we included all characteristics with either global p<0·05 or any level with p<0·001 vs reference, and then used backward elimination (exit p=0·05) to identify a final “main model”. We used a similar process on the behavioural variables, also adjusting for variables identified from the main screen, regardless of significance. We categorised screening variables into five broad groups dependent on persistence of effects (details in **Supplementary Methods**).

### Sensitivity Analyses

To assess the impact of small numbers of positives in some fortnights on power, we repeated the process using 28-day periods. Given logistic regression can have higher bias and variability with low rates, and hence lose accuracy and precision,^20^ we also compared the core variables effect estimates with those from ridge regression (see **Supplementary Results**).

## Results

Analyses included 4,091,537 RT-PCR results from nose and throat swabs from 482,677 individuals in 240,490 households from 19^th^ July 2020-17^th^ July 2021. 29,903 (0·7%) swabs were positive. Overall, the median (IQR) age was 52 years (33-66), 300,208 (7%) visits occurred in those reporting non-white ethnicity, 2,165,833 (53%) in females, 1,463,624 (36%) in major urban areas and 1,746,530 (43%) in urban cities/towns, most (1,735,618, 42%) in two-person households, and with a median deprivation percentile of 60 (34-81) (1=most deprived, 100=least deprived) (**Table 1**; screened variables in **Supplementary Table 1A,1B**). The highest positivity was 1·9% (95% CI 1·9-2·0%) 20^th^ December-2^nd^ January 2020, and the lowest 0·05% (0·03-0·08%) 2^nd^-15^th^ August 2020 (**Supplementary Figure 2A**). Numbers within each fortnight increased as the study expanded from August-October 2020,^21^ from 32,184 participants 19^th^ July-1^st^ August 2020 to a median 173,054 (IQR 168,171-195,031) from 27^th^ September 2020 onwards (**Supplementary Figure 3**).

**Table 1:** Characteristics of the core variables for visits included in analysis

| Characteristic | Positive, n (%) or median (IQR) | Negative, n (%) or median (IQR) | Total, n (%) or median (IQR) |
| --- | --- | --- | --- |
| **Age (years)** | 43 (23, 58) | 52 (33, 66) | 52 (33, 66) |
| **Sex** |  |  |  |
| Male | 14,405 (48) | 1,911,299 (47) | 1,925,704 (47) |
| Female | 15,498 (52) | 2,150,335 (53) | 2,165,833 (53) |
| **Ethnicity** |  |  |  |
| White | 26,702 (89) | 3,764,627 (93) | 3,791,329 (93) |
| Non-White | 3,201 (11) | 297,007 (7) | 300,208 (7) |
| **Deprivation percentile** | 54 (29, 78) | 60 (34, 81) | 60 (34, 81) |
| **Household (HH) size** |  |  |  |
| One | 3,842 (13) | 675,623 (17) | 679,465 (17) |
| Two | 10,124 (34) | 1,725,494 (42) | 1,735,618 (42) |
| Three | 5,797 (19) | 657,828 (16) | 663,625 (16) |
| Four | 6,639 (22) | 686,036 (17) | 692,675 (17) |
| Five or more | 3,501 (12) | 316,653 (8) | 320,154 (8) |
| **Multigenerational HH** |  |  |  |
| No | 27,311 (91) | 3,796,655 (93) | 3,823,966 (93) |
| Yes | 2,592 (9) | 264,979 (7) | 267,571 (7) |
| **Rural/urban classification** |  |  |  |
| Major urban area | 14,044 (47) | 1,449,580 (36) | 1,463,624 (36) |
| Urban city/town | 11,425 (38) | 1,735,105 (43) | 1,746,530 (43) |
| Rural town | 2,445 (8) | 435,296 (11) | 437,741 (11) |
| Rural village | 1,989 (7) | 441,653 (11) | 443,642 (11) |
| **Region** |  |  |  |
| London | 6,498 (22) | 698,608 (17) | 705,106 (17) |
| North West England | 5,077 (17) | 477,380 (12) | 482,457 (12) |
| North East England | 1,390 (5) | 156,119 (4) | 157,509 (4) |
| Yorkshire | 2,861 (10) | 343,353 (8) | 346,214 (8) |
| West Midlands | 2,266 (8) | 311,661 (8) | 313,927 (8) |
| East Midlands | 1,893 (6) | 264,293 (7) | 266,186 (7) |
| South East England | 2,986 (10) | 531,594 (13) | 534,580 (13) |
| South West England | 1,332 (4) | 320,869 (8) | 322,201 (8) |
| East England | 2,425 (8) | 405,304 (10) | 407,729 (10) |
| Northern Ireland | 665 (2) | 106,660 (3) | 107,325 (3) |
| Wales | 969 (3) | 179,900 (4) | 180,869 (4) |
| Scotland | 1,541 (5) | 265,893 (7) | 267,434 (7) |

Note: for deprivation percentile, 1=most deprived, 100=least deprived. Multigenerational household defined as households including individuals aged school year 11 or younger AND school year 12 to age 49 AND aged 50+

### Core model

From 19^th^ July-1^st^ August 2020, we found no evidence that any core variable was associated with positivity, potentially related to power given both low positivity (0·08% [95% CI 0·06-0·12%]) and sample size (32,184 swabs, 27 positive). The first characteristic associated with positivity was ethnicity, the only characteristic associated with positivity in the fortnights between 2^nd^-29^th^ August 2020 (**Figure 1A**), with 3·3 (1·1-10·0; p-value=0·034) and 3·5 (1·5-7·9; p-value=0·003) higher odds of positivity in those of non-white ethnicity, respectively.

**Figure 1A:**
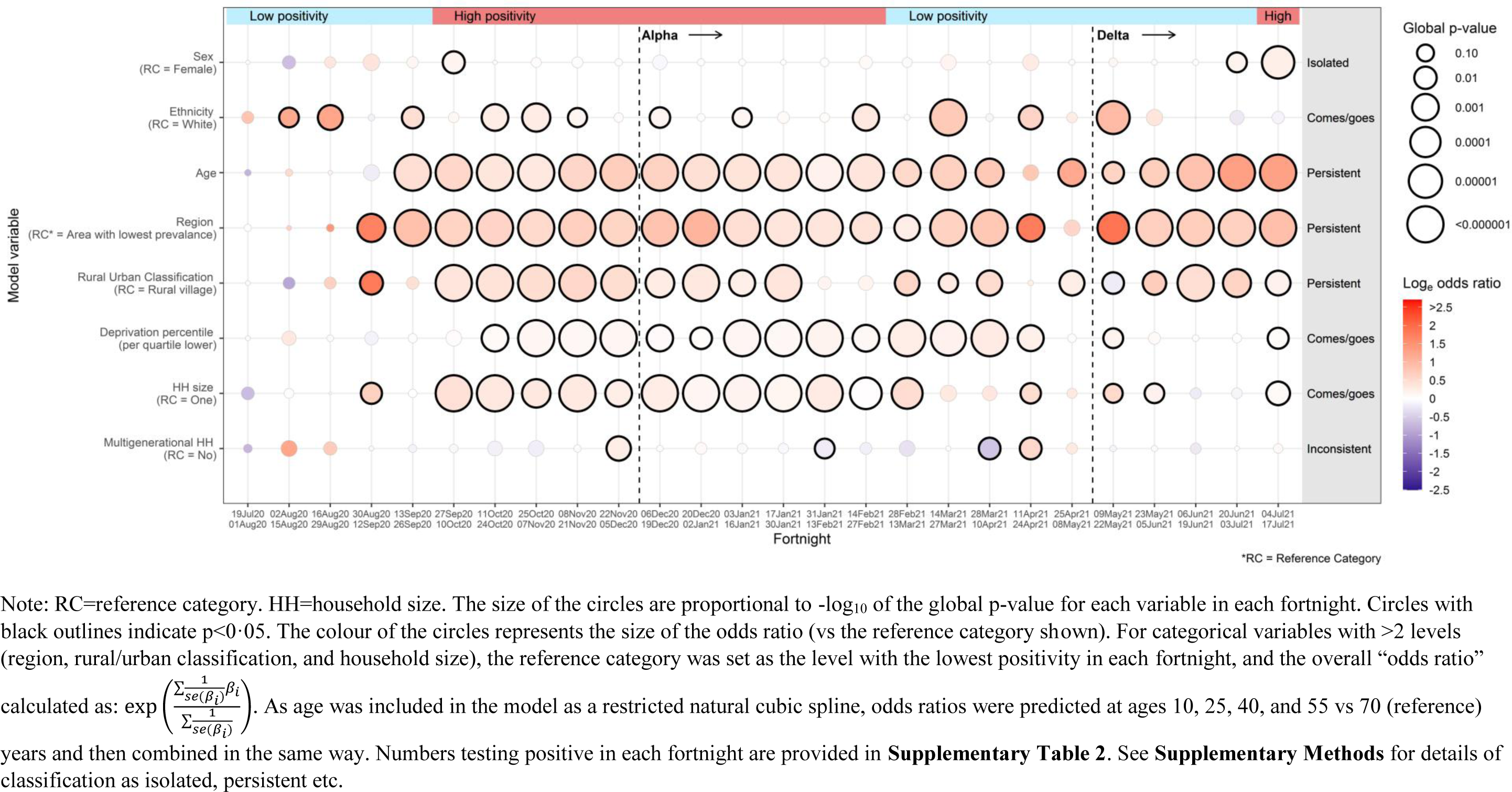
**Overall effects of the 8 core variables across the 52 week study period**

As positivity began to increase early September 2020, geographical region, rural/urban classification, and household size became independently associated with positivity, with odds of positivity highest in Wales, Northern Ireland, and northern English regions, in more urban areas, and those living in larger households (**Figure 1B**). For most subsequent fortnights, evidence of higher positivity persisted in participants living in more urban areas, and larger households.

**Figure 1B:**
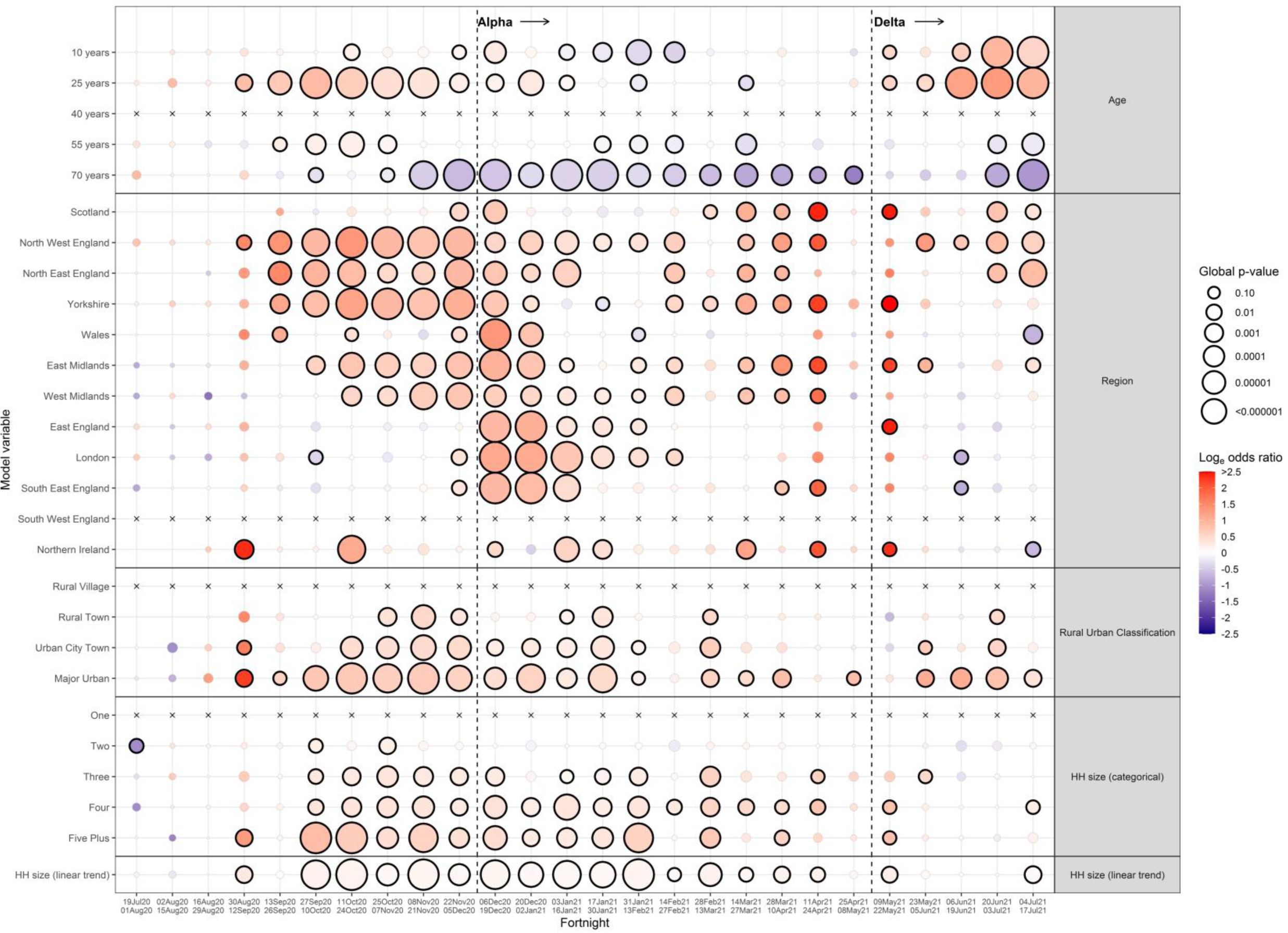
**Effects of the individual levels of the 8 core variables across the 52 week study period.**

As positivity rates rose further through October 2020, age and deprivation became associated with positivity, with rates highest in those 16-30y, and living in more deprived areas. Positivity was also heavily concentrated in northern and then midland English regions until 21^st^ November 2020. From 22^nd^ November, positivity increased overall, particularly in southern England, with higher odds of positivity in London, East, and South East England, reflecting the rise of the Alpha variant.^22^ Age remained strongly associated with positivity, but with less excess risk at younger ages, and instead decreased odds of positivity in those over 60y (**Figure 1B, Figure 2**). This lower risk in older individuals persisted for most subsequent fortnights. During February-May 2021, as positivity decreased, associations between positivity and age, region, and deprivation persisted, but their strength attenuated. As positivity rose during 17^th^ May-17^th^ July 2021, reflecting the rise of the Delta variant^23^ and major sporting events, sex was associated with positivity in two consecutive fortnights for the first time in the study, with higher odds in males compared with females. Age again became strongly associated, with a large peak in those aged 16-30y (**Figure 2**).

**Figure 2:**
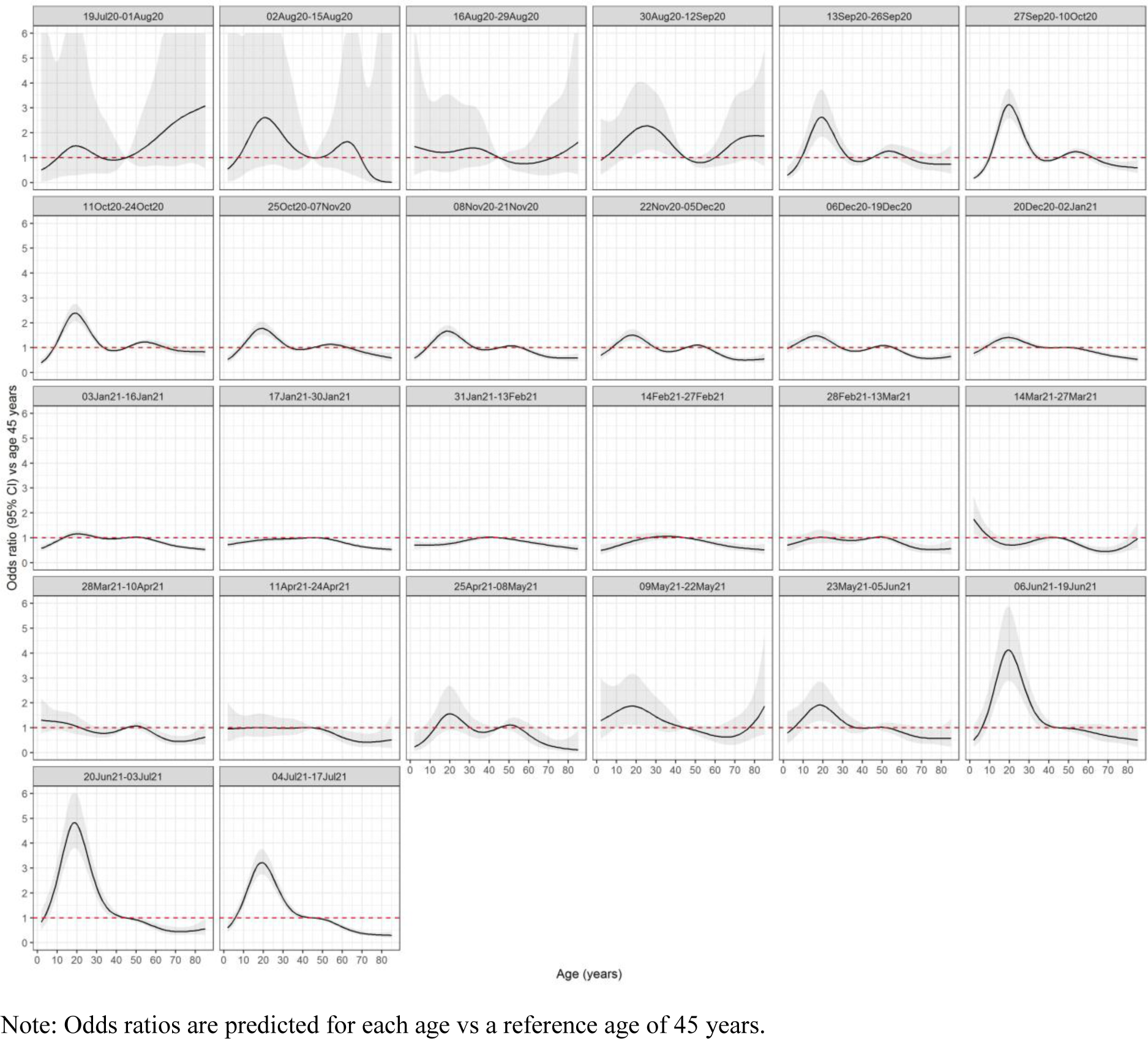
**Adjusted effect of age (years) on positivity over the 52 week study period.**

Few interactions between core variables were significant at the p=0·001 threshold, with no evidence of the same significant interactions in any consecutive fortnight (**Supplementary Figure 4**). For model comparability, none were therefore included in any fortnight for screening other variables.

### Screening process

As positivity increased, the screening process identified more variables and at a greater significance than expected by chance (**Figure 3**; **Figure 4A**). Contact with anyone who had recently had COVID- 19, currently self-isolating and thinking one had had COVID-19 recently, strongly and consistently predicted higher positivity. As these characteristics are potential mediators of effects of other factors, they were not considered further.

**Figure 3:**
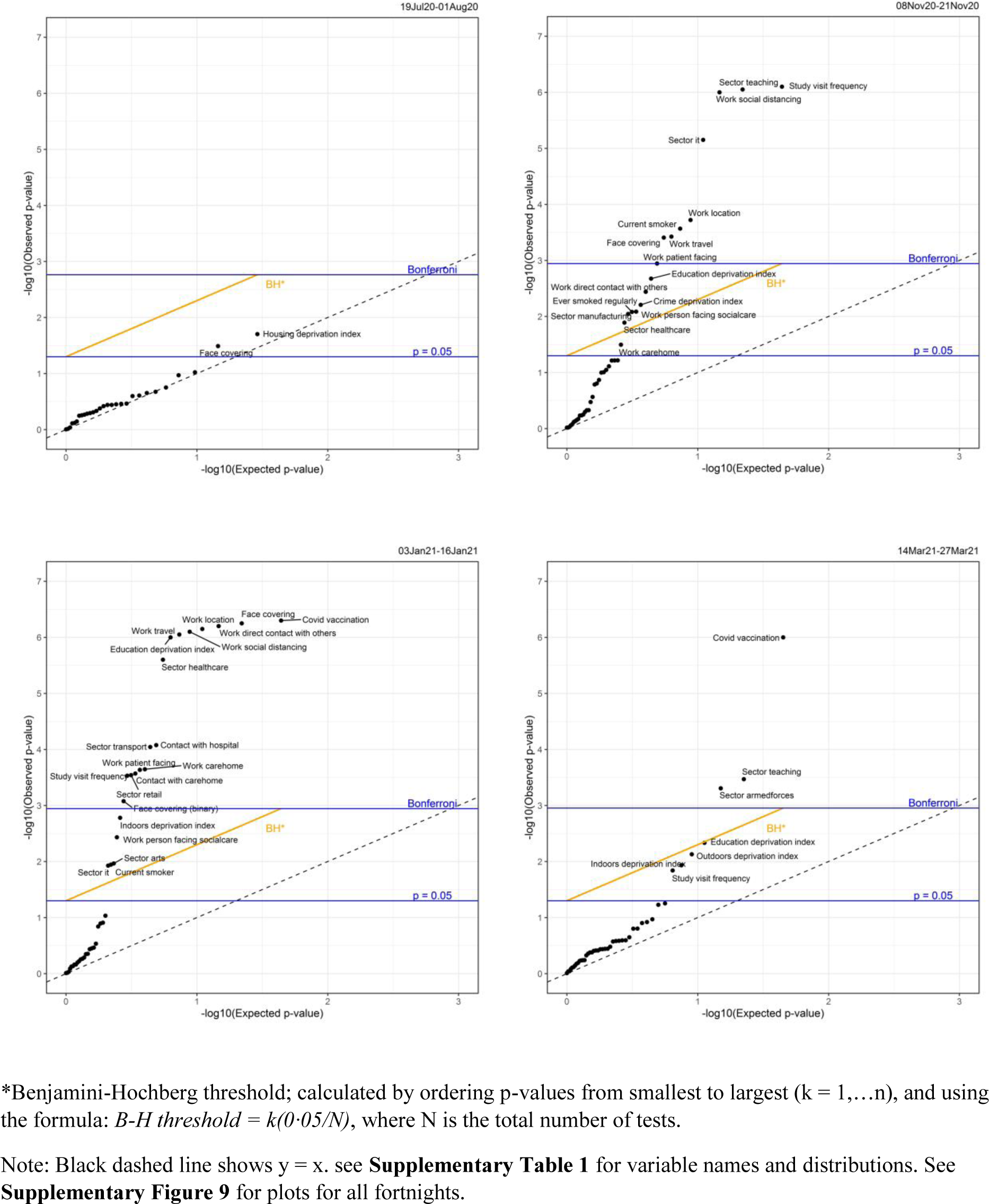
**Global heterogeneity p-values per factor from the screening process over 4 specific fortnights**

**Figure 4A:**
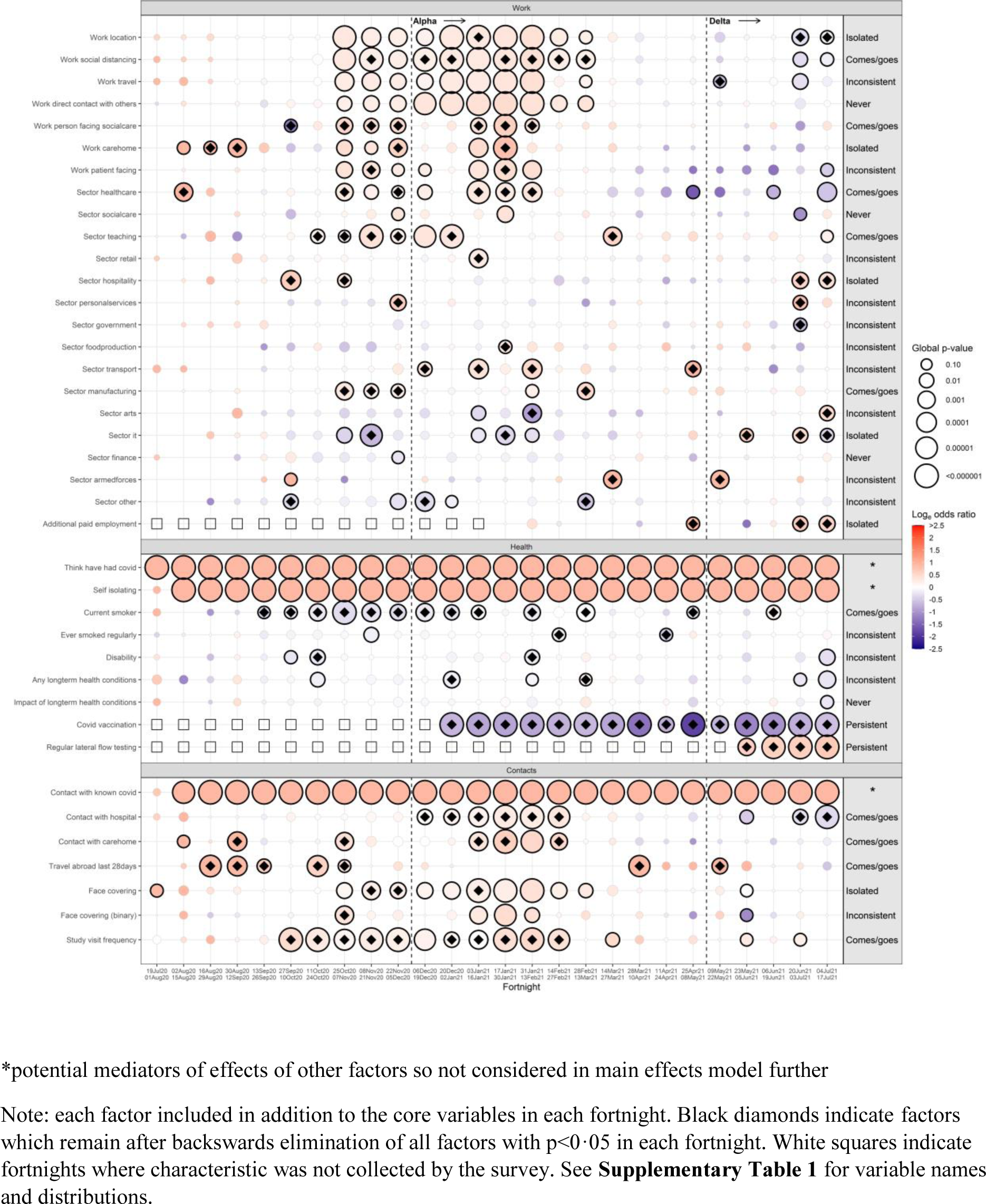
**Overall effects of additional factors from the screening process, adjusted for the core variables, over the 52 week study period**

Work and employment were significantly associated with positivity throughout the study. Initially from 2^nd^ August-12^th^ September 2020, there was independently higher positivity for those working in care/nursing homes or patient-facing healthcare roles (**Figure 4A**). This effect returned from 25^th^ October onwards, along with increased odds in those reporting working in healthcare sectors and specifically in person-facing social-care roles. From 25^th^ October 2020-27^th^ March 2021, we consistently observed higher positivity in those working outside compared with from home, with risk increasing as social distancing in the workplace became more difficult. Increased risk was also associated with all modes of travel to work (foot/bike, car/taxi, train/bus), compared with those not travelling to work (**Figure 4B**), with highest odds for car/taxi, then train/bus then foot/bike. Higher positivity was also observed in the teaching work sector during October/November 2020, while those working in IT had consistently lower odds (**Figure 4A**).

**Figure 4B.**
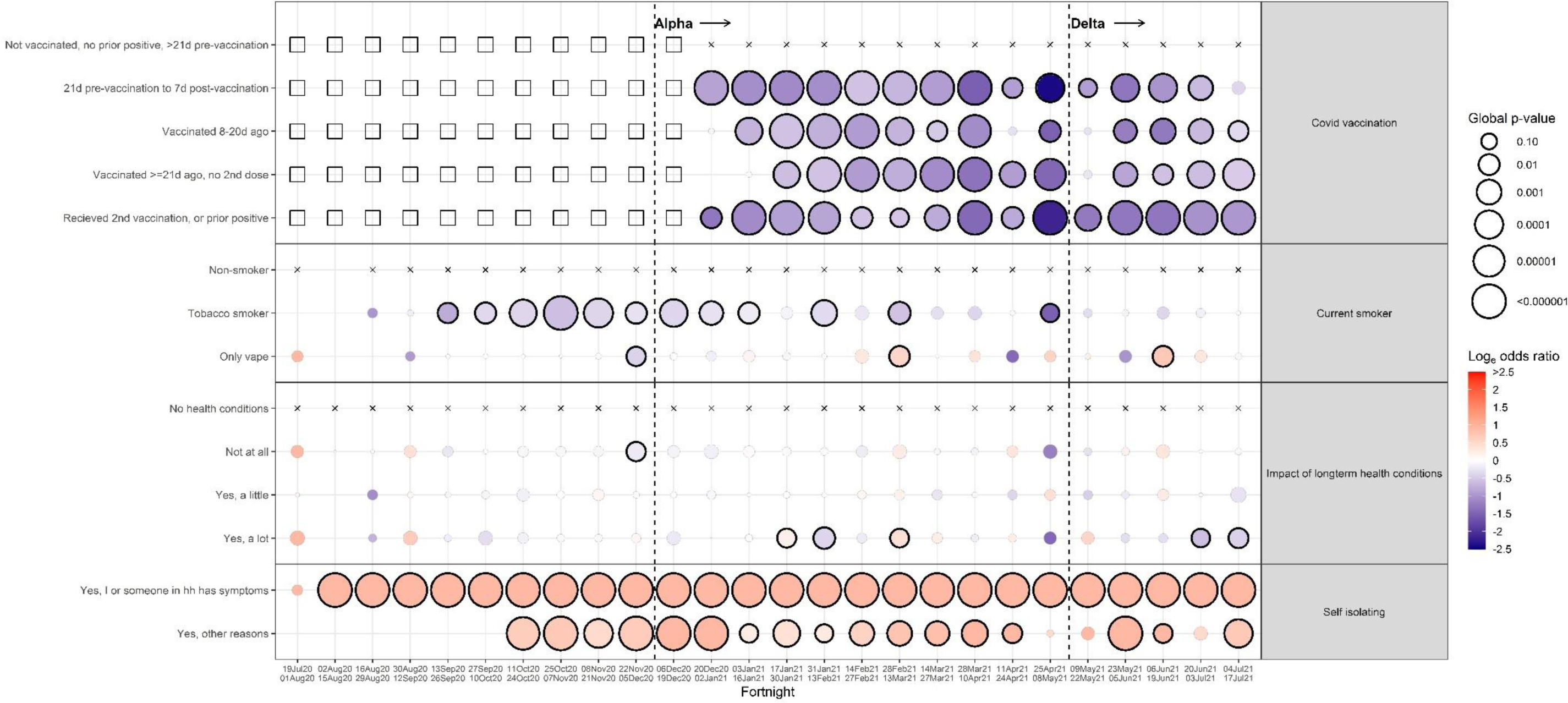
**Effects of individual levels of factors from the screening process, adjusted for the core variables, over the 52 week study period Health status**

From 16^th^ August-7^th^ November 2020, positivity was consistently higher in those who had travelled abroad in the last 28 days. This effect returned during 28^th^ March-12^th^ April 2021 and 9^th^-22^nd^ May 2021. Contact with hospital and care homes increased odds of positivity, particularly from 3^rd^ January-27^th^ February 2021, when positivity rates were very high due to Alpha. From 27^th^ September 2020-27^th^ February 2021 (when positivity was consistently >0·3%), participants were more likely to test positive on enrolment visits (**Figure 4B)**, most likely reflecting identification of longer-term PCR-positives at these visits.

Health-related variables varied in importance. Notably, there was no evidence of association between long-term health conditions and positivity. From 13^th^ September 2020-13^th^ March 2021, we consistently saw lower positivity in those who smoked tobacco products, compared with non-smokers. From 20^th^ December 2020, we observed a very strong effect of COVID-19 vaccination, with lower positivity in those vaccinated, compared with unvaccinated (**Figure 4B**). Deprivation components and living environment characteristics (available only for England) had little impact on positivity after adjusting for overall deprivation index and household size from the core model, likely due to high correlations between individual components with overall deprivation (**Supplementary Table 3; Supplementary Figure 5; Supplementary Results**).

Independently to the core model, we observed higher odds of positivity with increased social and physical contacts during periods when rates were high (**Figure 5; Supplementary Figure 6**). After also adjusting for variables identified from the main screening process and after backwards elimination, we observed higher odds of positivity with higher numbers of physical contacts with 18- 69 year olds between 20^th^ December 2020-13^th^ February 2021, and with higher numbers of physical contacts with those <18y between 14^th^ February 2021-27^th^ March 2021. As lockdown restrictions eased and Delta became prominent during 20^th^ June 2021-17^th^ July 2021, odds of positivity were higher in those with increasing time socialising outside home.

**Figure 5:**
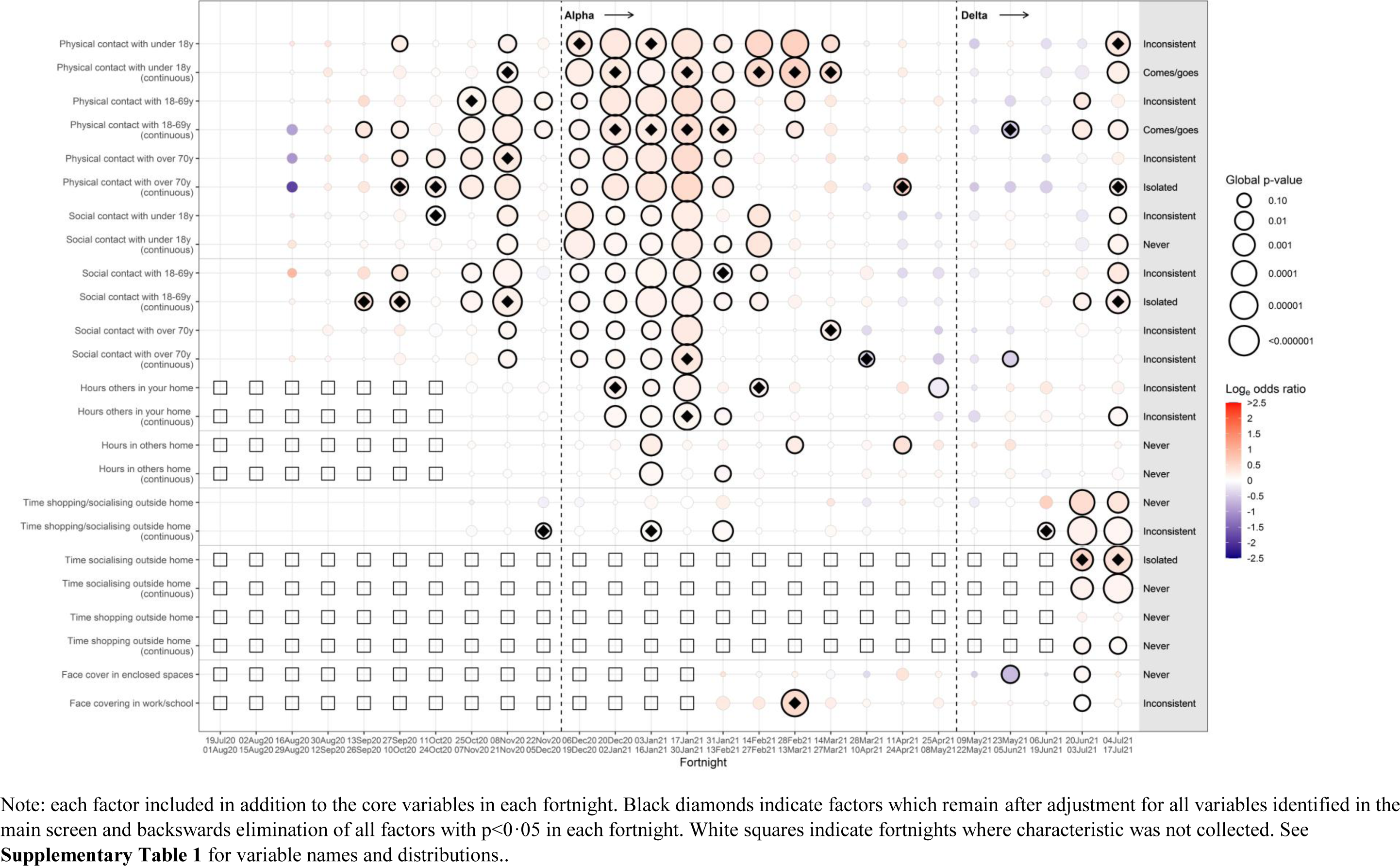
**Adjusted effects of behavioural variables from the screening process**

After backwards elimination, of the 71 variables screened (47 in the main screen, 13 variables in the behavioural screen with 24 parameterisations across the latter), two (3%) effects were persistent, 13 (18%) had effects which came and went, nine (13%) had effects isolated to only two consecutive fortnights, 30 (42%) were associated inconsistently in fortnights, and 17 (24%) were never associated.

### Sensitivity analysis

Similar key predictors of positivity were obtained using 28-day periods in the core model (**Supplementary Figures 7A,7B**,**8**). Notably, we saw a more consistent signal of higher positivity in non-white ethnicities from 11^th^ October 2020-27^th^ March 2021 (**Supplementary Figure 7A**), while this signal was more intermittent using fortnights (**Figure 1A**). We again did not see the same significant interactions in any consecutive 28-day periods (**Supplementary Figure 9A**). After backwards elimination, six interactions remained significant over five isolated 28-day periods (**Supplementary Figure 9B-G**). Three of these included household size, with a general pattern of stronger effects as household size increased in groups with higher positivity e.g. in younger ages (13^th^ September-10^th^ October 2020), non-white ethnicities (11^th^ October-7^th^ November 2020), and higher prevalence regions (6^th^ December 2020-2^nd^ January 2021). From 31^st^ January-27^th^ February 2021, compared with those living in non-multigenerational households, those of non-white ethnicities living in multigenerational households had increased odds of positivity, while those of white ethnicities had decreased odds.

Similar key associations were also identified from the screening process (**Supplementary Figure 10A, 10B**). Of the 45 consecutive occurrences of effects with p<0·05 in fortnights, 25 (56%) would have been detected later in 28-day periods, 14 (31%) at the same time, five (11%) earlier, and one (2%) never detected (**Supplementary Table 4**).

## Discussion

Over one year from 19^th^ July 2020-17^th^ July 2021, we estimated and summarised the key predictors of SARS-CoV-2 positivity in the UK, using a method designed to be run weekly in real-time to provide up-to-date information on changes in populations at increased risk. In the first fortnight from 19^th^ July-1^st^ August 2020, we had no evidence that any characteristic impacted positivity. As positivity rose through September-November 2020, they were independently higher in those of younger ages, living in Northern areas of England, in major urban conurbations, in more deprived areas, and in larger households. Additionally, rates were higher in those who had recently travelled abroad, worked in healthcare roles, or worked outside of home. As positivity peaked December 2020-January 2021, while we still observed strong effects of living in urban areas and large households, there was a major shift in high positivity to more southern geographical regions (reflecting the emergence of Alpha), with risk no longer concentrated in younger ages. Those working outside of home and in healthcare roles still had higher risk. As the national vaccine programme rolled out from December 2020, we saw large reductions in positivity in vaccinated individuals. From February-May 2021 as rates decreased, the impact of work on positivity decreased, while the effect of vaccination remained. As the Delta variant became prominent and positivity rates rose mid-May through July 2021, we observed higher odds of positivity in younger ages, in men, and in those not yet vaccinated.

The screening process demonstrated here has several limitations. First, low event numbers and smaller sample sizes reduce statistical power, reducing the chance of detecting true associations (false-negatives) and increasing the likelihood that the magnitude of “true” effects are inflated (false-positives).^24^ Increased statistical power using 28-day periods rather than fortnights more consistently detected associations with ethnicity in the core model and found more evidence of interactions. The screening process, however, detected the same characteristics using both time-periods, with earlier detection in most cases using fortnights. As there were no major differences and we aimed to identify associations most relevant to current positivity, the benefit of more regular estimates may outweigh the power gained from evaluating longer time-frames, although this will depend on event numbers.

When events numbers are low, logistic regression can be biased and/or imprecise.^25, 26^ Sensitivity analyses using penalised regression techniques showed most coefficients were within the logistic regression confidence intervals, suggesting that, while there was some attenuation of estimates, for example for geographical regions in a few fortnights, the logistic regression models were not substantially overfitting.

Multiple testing is an unavoidable limitation of our screening process. Doing many multiple independent tests increases the risk of false-positives;^27^ however, a priori the questionnaire was based on potential risk factors so the “correct” degree of adjustment is unclear. We therefore used Q-Q plots with Bonferroni and Benjamini-Hochberg adjustments to monitor the potential for false-positives, rather than as strict thresholds.^28, 29^ Even using stricter Bonferroni criteria, many screening variables were associated with positivity. Considering sex as a “negative control” (no effect expected), we only found an association in one of 24 fortnights before 20^th^ June 2021. The consistent association between sex and positivity from 20^th^ June-17^th^ July 2021 coincided with the European Football Championship, thus plausibly reflecting changes in social behaviour by sex, as observed elsewhere.^30^ Our results suggest more emphasis should be placed on effects that appear at least twice, interpreting effects that are inconsistent or appear sporadically with caution.

The underpinning design, namely a large community-based survey including randomly selected private households, is a major study strength. Participants being regularly asked about behaviours, work, and health status provided a rich opportunity to identify associations between positivity and many important demographic and behavioural characteristics. As participants were tested regardless of symptoms, characteristics could be assessed in an unbiased population, thus avoiding selection bias through only observing those choosing to take a COVID-19 test, for example, in the England national testing programme^31^ or through presenting to hospital with severe disease.

The study design also had limitations, particularly with individuals tested initially at weekly and then monthly visits. As fragments of virus can be detectable in the respiratory tract long after onset of infection, positives included in our outcome include both new infections and lingering PCR-positivity. Associations from the screening process may therefore not necessarily be related to new infections.

Whilst we could have grouped positive tests into “episodes”, for example, considering only the first positive in 90-day periods,^32^ we chose to mirror other point-prevalence studies, such as REACT,^14^ also expecting that many characteristics would be reasonably stable over time and therefore even associations with ongoing PCR-positivity could still be relevant to the original infection. This may however dilute effects if participants with long carriage have different characteristics to those testing positive with new infections. Ongoing PCR-positivity may also reduce sensitivity to detect specific “at-risk” populations as new variants emerge.

In conclusion, the screening process presented could be a valuable tool in understanding the characteristics driving current SARS-CoV-2 positivity, allowing us to provide enhanced up-to-date understanding of the pandemic across the UK. Looking forward, this could be used to target public health messages to detected groups to increased uptake of symptomatic and asymptomatic testing. We are using this method weekly to monitor the third wave of COVID-19 in the UK.

## Data Availability

De-identified study data are available for access by accredited researchers in the ONS Secure Research Service (SRS) for accredited research purposes under part 5, chapter 5 of the Digital Economy Act 2017.

## Main Figures and Tables

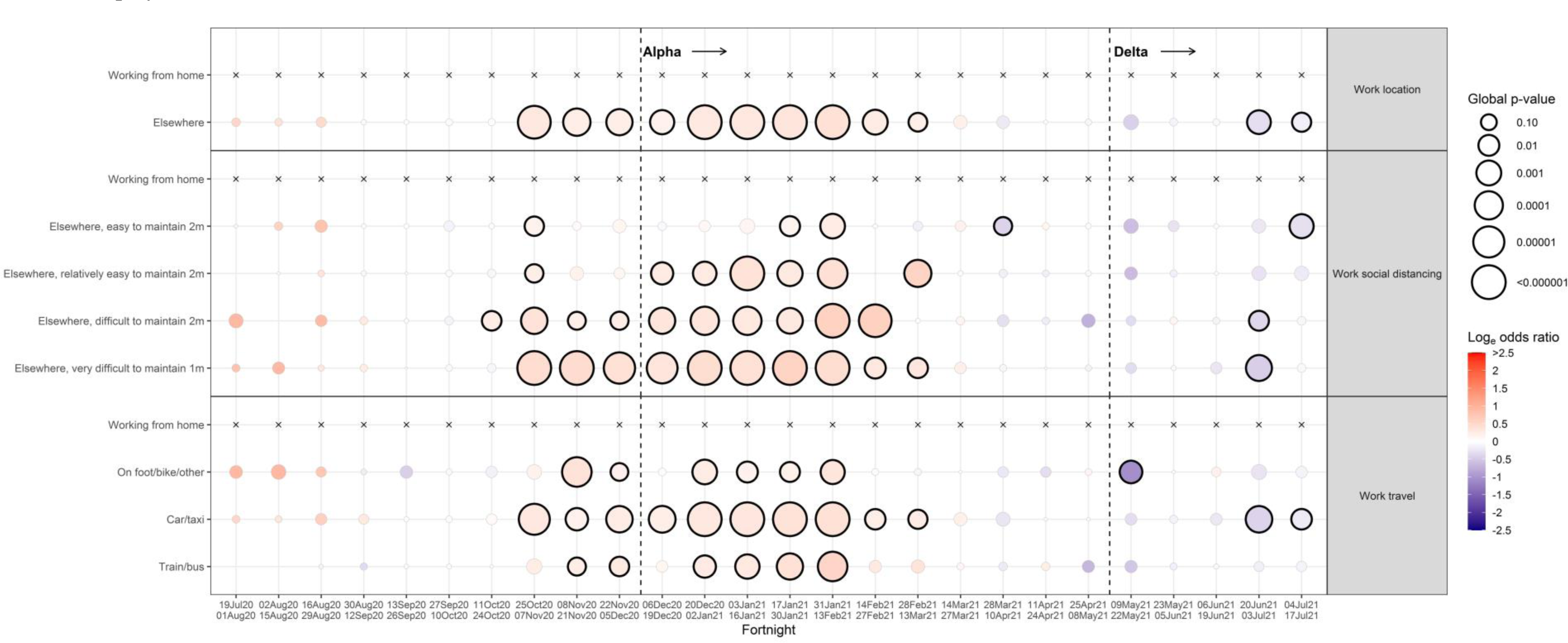
Work and employment

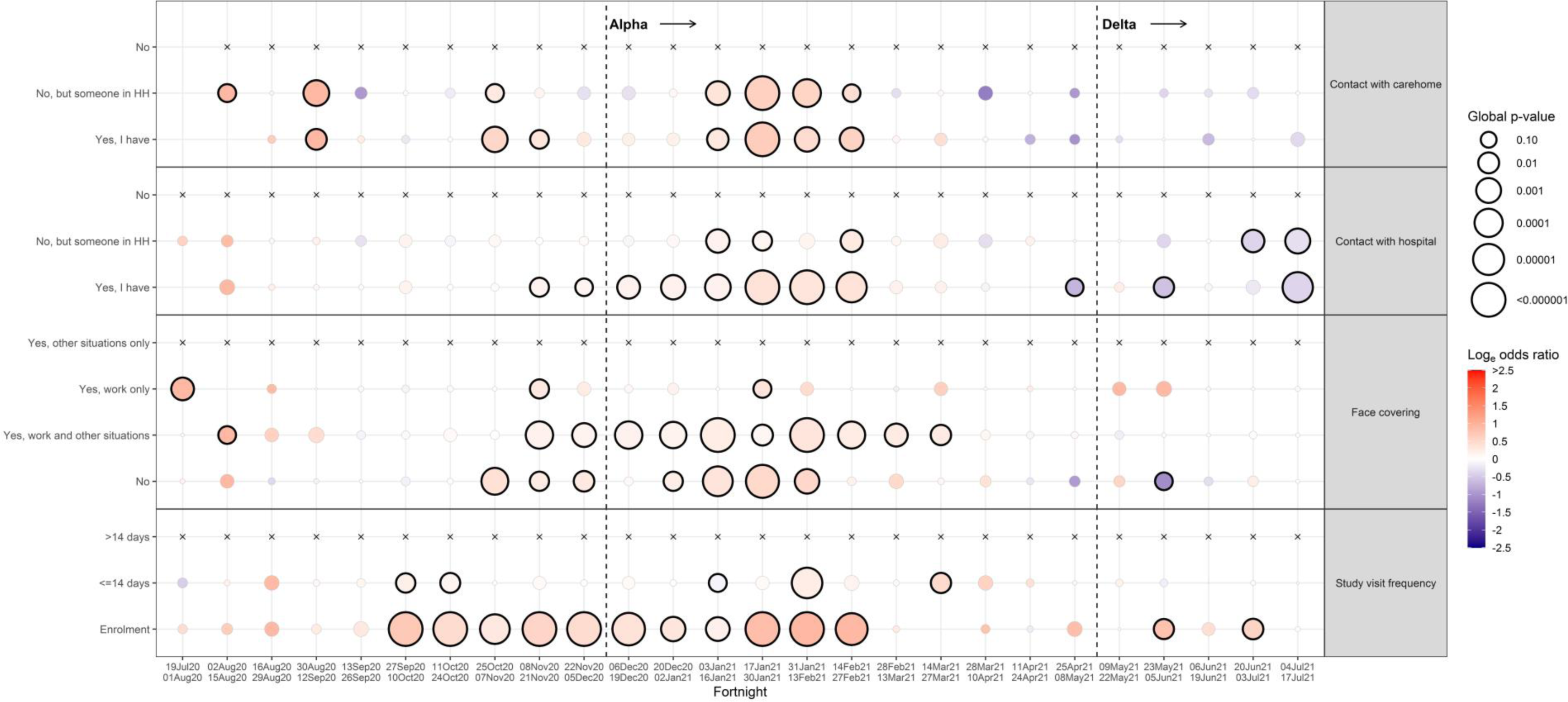
Contacts

## SUPPLEMENTARY MATERIAL

### Supplementary Methods

#### Laboratory testing

Swabs were couriered directly to the United Kingdom’s national Lighthouse laboratories (Glasgow (from 16 August 2020 onward) and the National Biocentre in Milton Keynes (from 26 April 2020 to 8 February 2021)) where samples were tested within the national testing program using identical methodology. The presence of three SARS-CoV-2 genes (ORF1ab and the genes transcribing nucleocapsid protein (N) and spike protein (S)) was identified using RT-PCR with the TaqPath RT-PCR COVID-19 kit (Thermo Fisher Scientific), analyzed using UgenTec FastFinder 3.300.5 (TaqMan 2019-nCoV Assay Kit V2 UK NHS ABI 7500 v2.1; UgenTec). The assay plugin contained an assay-specific algorithm and decision mechanism allowing conversion of the qualitative amplification assay raw data into test results with little manual intervention. Samples were called positive if either N or ORF1ab, or both, were detected. The S gene alone was not considered a reliable positive but could accompany other genes (that is, one, two or three gene positives).

### Variable and model specifications

#### Deprivation

Deprivation was assessed using the index of multiple deprivation (IMD) in England, a score based on lower layer super output areas with average population of 1500 people and incorporating seven domains to produce an overall relative measure of deprivation (income, employment, education, skills and training, health and disability, crime, barriers to housing services and living environment) (https://www.gov.uk/government/statistics/english-indices-of-deprivation-2019). These sub-components were also assessed in the variable screening process, restricted to England. Equivalent scores were used in the other three countries comprising the UK^1–4^. Each country’s scores were converted to a within country percentile.

#### Age

Age was including in the model as a natural cubic spline with 4 internal knots at 20, 40, 60, 80th percentiles of unique ages, and boundary knots at 5th and 95th percentiles.

#### Vaccination status

Participants were asked about their vaccination status at visits, including the type, number of doses and date(s). Participants from England were also linked to administrative records from the National Immunisation Management Service (NIMS). We used records from NIMS where available. Otherwise, we used records from the survey, since linkage was periodic and NIMS does not contain information about vaccinations received abroad or in Northern Ireland, Scotland and Wales. Where records were available from both NIMS and the survey, agreement on type was 98% and agreement on dates was 95% within ±7 days.

#### Interactions

Interactions between household size and multigenerational households, and region and rural/urban classification were not considered as, by definition, all those living in multigenerational households had a household size of 3 or more, and not all regions included major urban conurbations.

#### Face covering variables

Prior to 18^th^ February, participants in the study were asked the following question regarding face coverings: “*Do you mainly wear any kind of face covering or mask when you are outside your home, because of COVID-19?*” with the options:

– “No”,
– “Yes, at work/school only”,
– “Yes in other situations only (including public transport, shops)”,
– “Yes, usually both at work/school and in other situations”
– “My face is already covered for other reasons (e.g. religious or cultural reasons)”

As of 18^th^ February this question was retired, and participants were instead asked the two following questions about face coverings: “*Do you wear any kind of face covering or mask when you are at work/your place of education, because of COVID19?”*, and “*Do you wear any kind of face covering or mask when you are in other enclosed public spaces, such as shops, or using public transport, because of COVID-19?*”, with the first options being either “Not going to place of work or education”, or “Not going to place of work or education”. This question caused similar issues with reverse causality as other behavioural questions, and hence these new questions were including in our behavioural screen, while the former question was included in the main screen.

#### Approximate categorisation of variable effects

We classified effects from each variable in both the core and screening model using the following broad categorisation:

- **Never:** The effect is never significant at a p<0·05 threshold in any fortnight
- **Inconsistent**: The variable is significant at a p<0·05 threshold in at least one fortnight, but never in with an odds ratio in a consistent direction in any consecutive fortnights
- **Isolated**: The variable is significant at a p<0·05 threshold in two consecutive fortnight at most once, and “never consecutive” at all other times
- **Comes/goes**: The variable is significant at a p<0·05 threshold in three or more consecutive fortnights, or two consecutive fortnights at least twice, and is not significant with a gap of at least three fortnights, or two gaps of two fortnights, if the effect appears again.
- **Persistent**: The variable is significant at a p<0·05 threshold for the entire period after the first significant fortnight, with no more than one gap of two fortnights separating consistency of the effect.

## Supplementary Results

While the deprivation score component reflecting education was consistently associated with positivity, as this effect was in the same direction as the main deprivation score in the core model, and was only available in England, it was not considered further (**Supplementary Figure 4)**.

### Ridge regression

We found 43 of the 692 (6%) coefficients from the core models produced from ridge regression did not fall within the 95% of the equivalent coefficients obtained from logistic regression (**Supplementary Figure 10**). Of these, the majority (38 coefficients; 88%) were effects of geographical region. These were mostly in the first fortnights of the study period when event rates and sample size was smallest, and also during December 2020, where we observed strong regional effects due to the rise of the Alpha variant in the Southern regions of England. Many of the inconsistencies within geographical region occurred within the same fortnight i.e. either none or all of the effect estimates for geographic regions were within the confidence intervals.

The differences observed between coefficients in December 2020 while the Alpha variant was rising suggest that the ridge regression penalised early signal for the regional effect, while logistic regression models picked this up. While often challenging to distinguish between signal and noise, through triangulation with other data sources, the regional effects observed in logistic regression model were accurate and representative of rises in Alpha variant in London and the South East, while ridge regression missed this effect, hence justifying our choice of method.

## Supplementary Tables

**Supplementary Table 1A:**
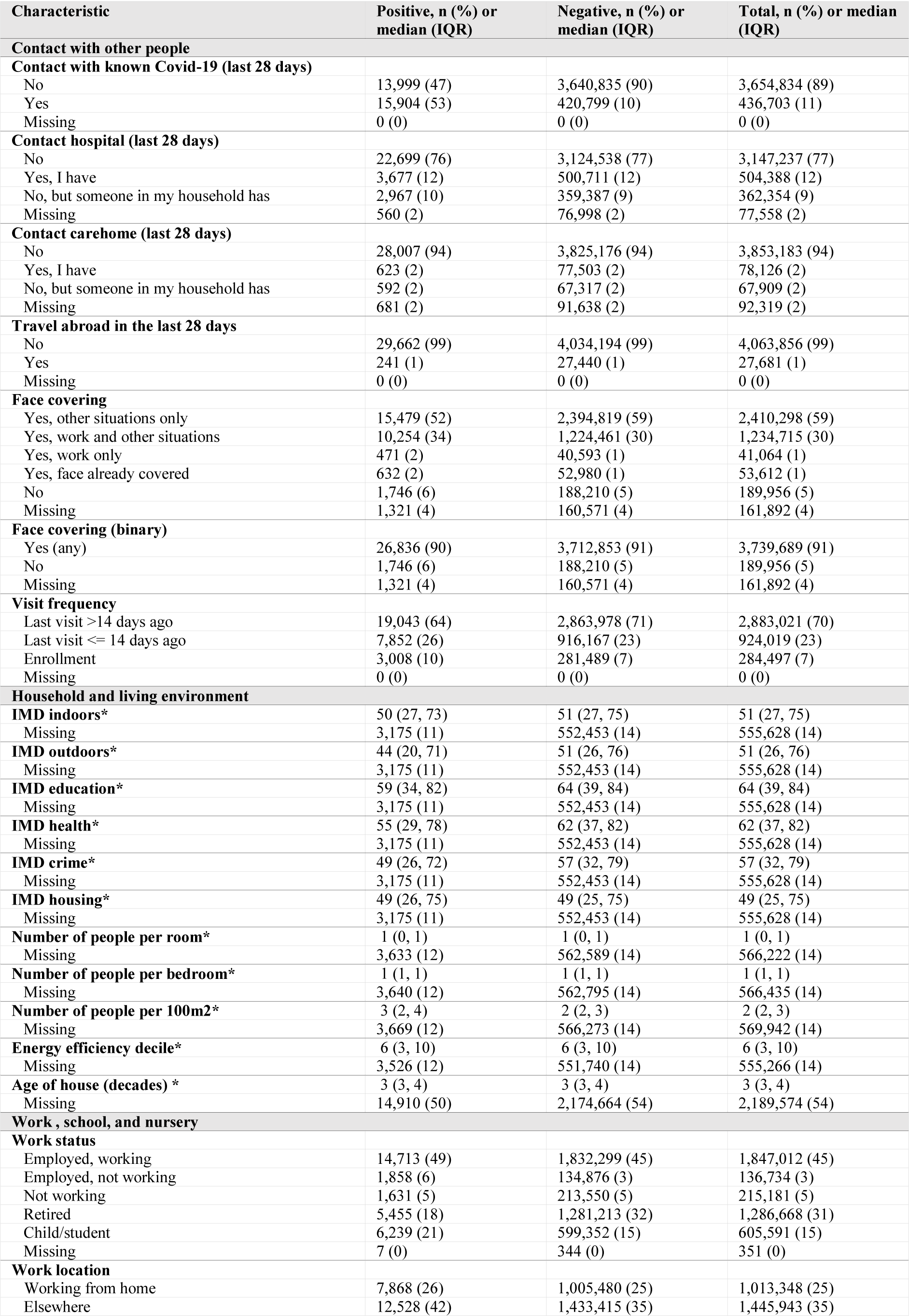
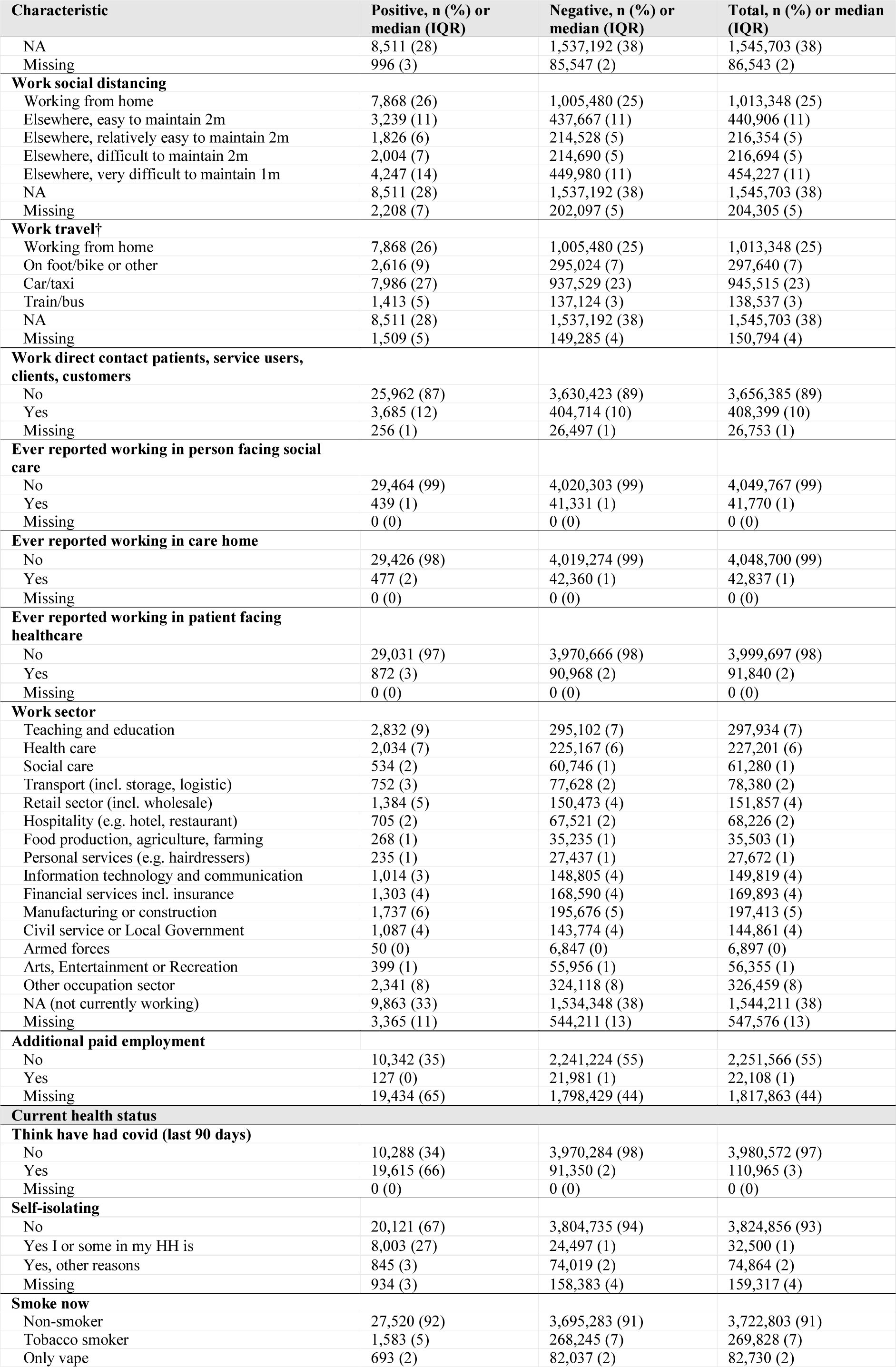
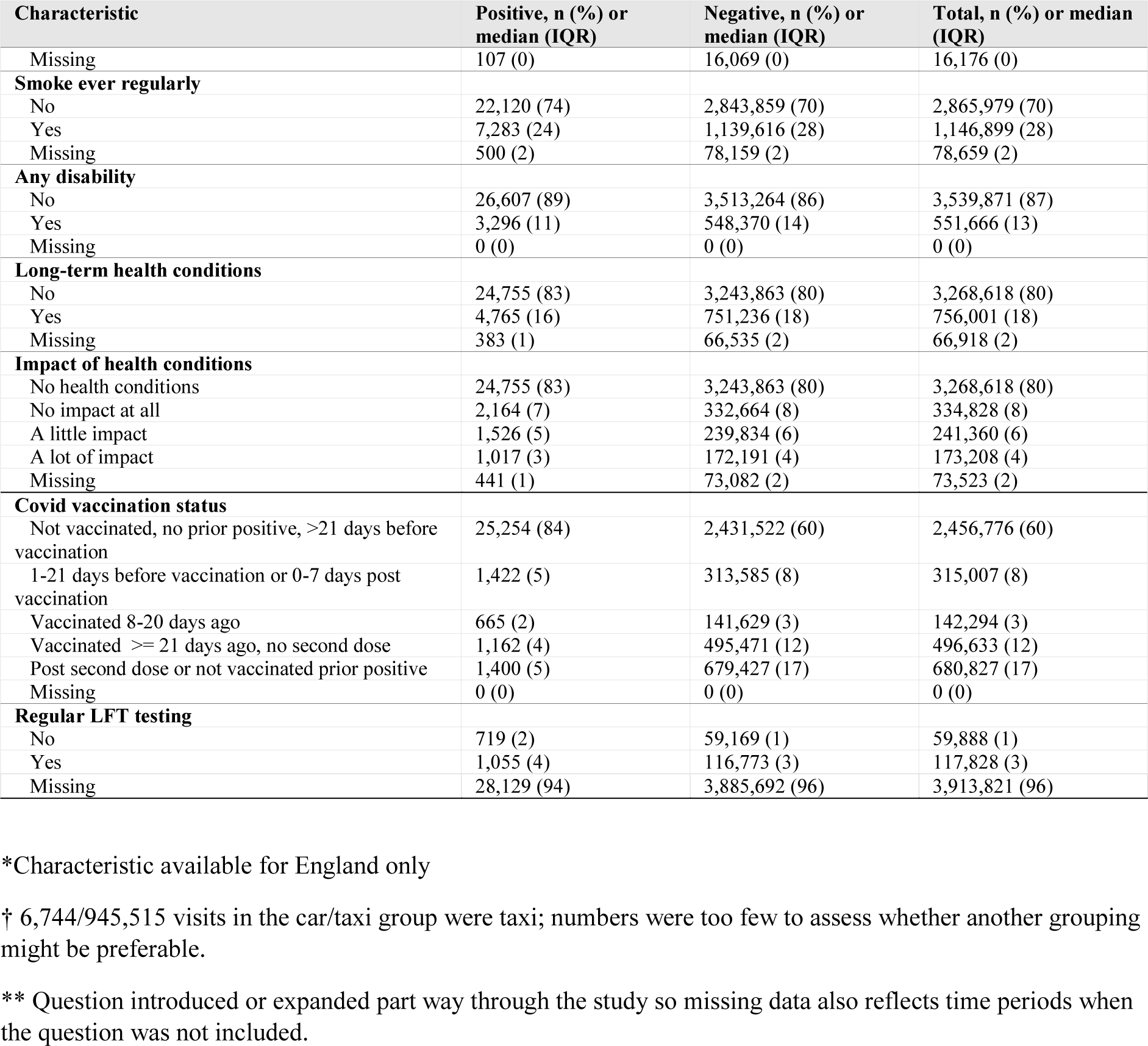
Characteristics of screening variables for visits included in the main screening process

**Supplementary Table 1B.** Characteristics of screening variables for visits included in the behaviour screening process (B)

| Characteristic† | Positive, n (%) or median (IQR) | Negative, n (%) or median (IQR) | Total, n (%) or median (IQR) |
| --- | --- | --- | --- |
| **Number of physical contacts aged <18** |  |  |  |
| 0 | 11,898 (40) | 2,160,467 (53) | 2,172,365 (53) |
| 1-5 | 4,146 (14) | 608,127 (15) | 612,273 (15) |
| 6-10 | 675 (2) | 71,849 (2) | 725,24 (2) |
| 11-20 | 2,294 (8) | 206,076 (5) | 208,370 (5) |
| 21 or more | 10,890 (36) | 1,015,115 (25) | 1,026,005 (25) |
| Missing | 11,898 (40) | 2,160,467 (53) | 2,172,365 (53) |
| **Number of physical contacts aged 18-69** |  |  |  |
| 0 | 10,031 (34) | 1,848,906 (46) | 1,858,937 (45) |
| 1-5 | 6,487 (22) | 950,800 (23) | 957,287 (23) |
| 6-10 | 1,233 (4) | 128,817 (3) | 130,050 (3) |
| 11-20 | 1,269 (4) | 119,866 (3) | 121,135 (3) |
| 21 or more | 10,883 (36) | 1,013,245 (25) | 1,024,128 (25) |
| Missing | 10,031 (34) | 1,848,906 (46) | 1,858,937 (45) |
| **Number of physical contacts aged >=70** |  |  |  |
| 0 | 15,293 (51) | 2,530,655 (62) | 2,545,948 (62) |
| 1-5 | 3,034 (10) | 449,008 (11) | 452,042 (11) |
| 6-10 | 205 (1) | 22,434 (1) | 22,639 (1) |
| 11-20 | 423 (1) | 41,165 (1) | 41,588 (1) |
| 21 or more | 10,948 (37) | 1,018,372 (25) | 1,029,320 (25) |
| Missing | 15,293 (51) | 2,530,655 (62) | 2,545,948 (62) |
| **Number of social contacts aged <18** |  |  |  |
| 0 | 12,138 (41) | 1,935,681 (48) | 1,947,819 (48) |
| 1-5 | 4,797 (16) | 835,491 (21) | 840,288 (21) |
| 6-10 | 696 (2) | 106,921 (3) | 107,617 (3) |
| 11-20 | 1,294 (4) | 163,396 (4) | 164,690 (4) |
| 21 or more | 10,978 (37) | 1,020,145 (25) | 1,031,123 (25) |
| Missing | 12,138 (41) | 1,935,681 (48) | 1,947,819 (48) |
| **Number of social contacts aged 18-69** |  |  |  |
| 0 | 4,243 (14) | 803,071 (20) | 807,314 (20) |
| 1-5 | 6,351 (21) | 1,191,642 (29) | 1,197,993 (29) |
| 6-10 | 3,033 (10) | 425,739 (10) | 428,772 (10) |
| 11-20 | 5,385 (18) | 628,365 (15) | 633,750 (15) |
| 21 or more | 10,891 (36) | 1,012,817 (25) | 1,023,708 (25) |
| Missing | 4,243 (14) | 803,071 (20) | 807,314 (20) |
| **Number of social contacts aged >=70** |  |  |  |
| 0 | 12,138 (41) | 1,935,681 (48) | 1,947,819 (48) |
| 1-5 | 4,797 (16) | 835,491 (21) | 840,288 (21) |
| 6-10 | 696 (2) | 106,921 (3) | 107,617 (3) |
| 11-20 | 1,294 (4) | 163,396 (4) | 164,690 (4) |
| 21 or more | 10,978 (37) | 1,020,145 (25) | 1,031,123 (25) |
| Missing | 12,138 (41) | 1,935,681 (48) | 1,947,819 (48) |
| **Outside socialising times** |  |  |  |
| None | 409 (1) | 80,290 (2) | 80,699 (2) |
| Once | 345 (1) | 52,733 (1) | 53,078 (1) |
| Twice | 208 (1) | 28,400 (1) | 28,608 (1) |
| Three times | 128 (0) | 14,056 (0) | 14,184 (0) |
| Four times | 54 (0) | 6,834 (0) | 6,888 (0) |
| Five times | 52 (0) | 4,194 (0) | 4,246 (0) |
| Six times | 19 (0) | 1,468 (0) | 1,487 (0) |
| Seven times or more | 43 (0) | 4,718 (0) | 4,761 (0) |
| Missing | 28,645 (96) | 3,868,941 (95) | 3,897,586 (95) |
| **Outside shopping only times** |  |  |  |
| None | 260 (1) | 32,514 (1) | 32,774 (1) |
| Once | 297 (1) | 47,098 (1) | 47,395 (1) |
| Twice | 291 (1) | 48,764 (1) | 49,055 (1) |
| Three times | 180 (1) | 30,207 (1) | 30,387 (1) |
| Four times | 84 (0) | 13,948 (0) | 14,032 (0) |
| Five times | 56 (0) | 7,835 (0) | 7,891 (0) |
| Six times | 14 (0) | 2,663 (0) | 2,677 (0) |
| Seven times or more | 76 (0) | 9,669 (0) | 9,745 (0) |
| Missing | 28,645 (96) | 3,868,936 (95) | 3,897,581 (95) |
| **Time spent shopping or socializing outside** |  |  |  |
| None | 3,180 (11) | 513,784 (13) | 516,964 (13) |
| Once | 3,687 (12) | 634,651 (16) | 638,338 (16) |
| Twice | 3,719 (12) | 602,006 (15) | 605,725 (15) |
| Three times | 2,236 (7) | 356,644 (9) | 358,880 (9) |
| Four times | 1,133 (4) | 180,386 (4) | 181,519 (4) |
| Five times | 737 (2) | 111,370 (3) | 112,107 (3) |
| Six times | 293 (1) | 44,966 (1) | 45,259 (1) |
| Seven times or more | 1,196 (4) | 177,612 (4) | 178,808 (4) |
| Missing | 13,722 (46) | 1,440,215 (35) | 1,453,937 (36) |
| **Hours spent in other's homes** |  |  |  |
| None | 11,597 (39) | 1,954,027 (48) | 1,965,624 (48) |
| Once | 2,619 (9) | 397,279 (10) | 399,898 (10) |
| Twice | 830 (3) | 125,436 (3) | 126,266 (3) |
| Three | 356 (1) | 49,845 (1) | 50,201 (1) |
| Four | 175 (1) | 23,312 (1) | 23,487 (1) |
| Five | 129 (0) | 19,331 (0) | 19,460 (0) |
| Six | 49 (0) | 6,566 (0) | 6,615 (0) |
| Seven or more | 261 (1) | 34,453 (1) | 34,714 (1) |
| Missing | 13,887 (46) | 1,451,385 (36) | 1,465,272 (36) |
| **Hours others spent in own home** |  |  |  |
| None | 10,753 (36) | 1,780,000 (44) | 1,790,753 (44) |
| Once | 2,906 (10) | 490,148 (12) | 493,054 (12) |
| Twice | 1,139 (4) | 171,857 (4) | 172,996 (4) |
| Three times | 502 (2) | 69,515 (2) | 70,017 (2) |
| Four times | 201 (1) | 32,278 (1) | 32,479 (1) |
| Five times | 180 (1) | 23,641 (1) | 23,821 (1) |
| Six times | 60 (0) | 7,818 (0) | 7,878 (0) |
| Seven times or more | 254 (1) | 32,741 (1) | 32,995 (1) |
| Missing | 13,908 (47) | 1,453,636 (36) | 1,467,544 (36) |
| **Face coverings (work/school)** |  |  |  |
| Not going to work/school | 1,491 (5) | 854,693 (21) | 856,184 (21) |
| Never | 1,190 (4) | 413,563 (10) | 414,753 (10) |
| Yes, sometimes | 433 (1) | 111,707 (3) | 112,140 (3) |
| Yes, always | 434 (1) | 124,250 (3) | 124,684 (3) |
| Face already covered | 26,355 (88) | 2,557,421 (63) | 2,583,776 (63) |
| Missing | 1,491 (5) | 854,693 (21) | 856,184 (21) |
| **Face coverings (other situations)** |  |  |  |
| Yes, always | 113 (0) | 49,684 (1) | 49,797 (1) |
| Yes, sometimes | 3,156 (11) | 1,376,289 (34) | 1,379,445 (34) |
| Face already covered | 120 (0) | 39,674 (1) | 39,794 (1) |
| Not going to enclosed public spaces | 181 (1) | 49,240 (1) | 49,421 (1) |
| Never | 26,333 (88) | 2,546,747 (63) | 2,573,080 (63) |
| Missing | 113 (0) | 49,684 (1) | 49,797 (1) |

† All characteristics except hours spent with someone else in one's own home per day relate to the past 7 days.

**Supplementary Table 2:** Count in each fortnight, including number not included in core model

| Fortnight | Positive visits, n (%) | Negative visits, n (%) | Total, n (%) | Negative visits excluded from core models*, n (% of negatives) |
| --- | --- | --- | --- | --- |
| 19Jul20-01Aug20 | 27 (0.1) | 32,157 (99.9) | 32,184 (100) | 4,074 (12.7) |
| 02Aug20-15Aug20 | 22 (0.1) | 43,073 (99.9) | 43,095 (100) | 86,72 (20.1) |
| 16Aug20-29Aug20 | 41 (0.1) | 57,895 (99.9) | 57,936 (100) | 0 (0.0) |
| 30Aug20-12Sep20 | 111 (0.1) | 76,276 (99.9) | 76,387 (100) | 0 (0.0) |
| 13Sep20-26Sep20 | 320 (0.3) | 116,467 (99.7) | 116,787 (100) | 0 (0.0) |
| 27Sep20-10Oct20 | 1,090 (0.6) | 171,298 (99.4) | 172,388 (100) | 0 (0.0) |
| 11Oct20-24Oct20 | 1,995 (1.0) | 194,123 (99.0) | 196,118 (100) | 0 (0.0) |
| 25Oct20-07Nov20 | 2,109 (1.2) | 169,735 (98.8) | 171,844 (100) | 0 (0.0) |
| 08Nov20-21Nov20 | 2,316 (1.2) | 192,715 (98.8) | 195,031 (100) | 0 (0.0) |
| 22Nov20-05Dec20 | 1,874 (1.0) | 192,534 (99.0) | 194,408 (100) | 0 (0.0) |
| 06Dec20-19Dec20 | 2,286 (1.2) | 190,313 (98.8) | 192,599 (100) | 0 (0.0) |
| 20Dec20-02Jan21 | 2,710 (1.9) | 136,703 (98.1) | 139,413 (100) | 0 (0.0) |
| 03Jan21-16Jan21 | 3,891 (1.9) | 198,116 (98.1) | 202,007 (100) | 0 (0.0) |
| 17Jan21-30Jan21 | 3,275 (1.7) | 194,157 (98.3) | 197,432 (100) | 0 (0.0) |
| 31Jan21-13Feb21 | 2,171 (1.0) | 205,148 (99.0) | 207,319 (100) | 0 (0.0) |
| 14Feb21-27Feb21 | 1,058 (0.5) | 196,410 (99.5) | 197,468 (100) | 0 (0.0) |
| 28Feb21-13Mar21 | 621 (0.3) | 193,549 (99.7) | 194,170 (100) | 0 (0.0) |
| 14Mar21-27Mar21 | 475 (0.3) | 173,734 (99.7) | 174,209 (100) | 0 (0.0) |
| 28Mar21-10Apr21 | 364 (0.2) | 169,692 (99.8) | 170,056 (100) | 0 (0.0) |
| 11Apr21-24Apr21 | 189 (0.1) | 164,958 (99.9) | 165,147 (100) | 0 (0.0) |
| 25Apr21-08May21 | 123 (0.1) | 172,931 (99.9) | 173,054 (100) | 0 (0.0) |
| 09May21-22May21 | 137 (0.1) | 164,249 (99.9) | 164,386 (100) | 0 (0.0) |
| 23May21-05Jun21 | 240 (0.1) | 160,888 (99.9) | 161,128 (100) | 0 (0.0) |
| 06Jun21-19Jun21 | 309 (0.2) | 167,862 (99.8) | 168,171 (100) | 0 (0.0) |
| 20Jun21-03Jul21 | 675 (0.4) | 159,246 (99.6) | 159,921 (100) | 0 (0.0) |
| 04Jul21-17Jul21 | 1,474 (0.9) | 167,405 (99.1) | 168,879 (100) | 0 (0.0) |

\* Negative visits were excluded in the two earliest fortnights due to perfect prediction

**Supplementary Table 3:** Summary of individuals IMD components with combined index

|  | Correlation with combined index | Proportion of score* |
| --- | --- | --- |
| Combined | 1 |  |
| Income | 0.93 | 22.5 |
| Employment | 0.90 | 22.5 |
| Education | 0.78 | 13.5 |
| Health | 0.81 | 13.5 |
| Crime | 0.68 | 9.3 |
| Housing | 0.18 | 9.3 |
| Indoors | 0.35 | 6.2 |
| Outdoors | 0.25 | 3.1 |
| Living environment (combination of “indoors” and “outdoors”) | 0.41 | 9.3 |

\*Taken from https://www.gov.uk/government/statistics/english-indices-of-deprivation-2019

**Supplementary Table 4:** Summary of p-values in 28-day periods for effects which occur in 2 or more consecutive fortnights

|  | Number of occurrences, n (%) [N=45] |
| --- | --- |
| Not detected in 28-day periods | 1 (2) |
| Same detection date | 14 (31) |
| Detected later in 28-day periods | 25 (56) |
| Detected earlier in 28-day periods | 5 (11) |

Note: The effect not detected in 28-day periods was work sector IT in the fortnight 20<sup>th</sup> June-3<sup>rd</sup> July. Variables which would have been detected earlier in 28-day periods (number of days earlier in brackets) are as follows: contact with hospital (14 days), work in a patient facing healthcare role (14 days), education deprivation index (14 days), sector health care (42 days), study visit frequency (56 days).
Additional to these earlier detections, for eight variables in ten 28-day periods, the effect had $p < 0.05$ in a 28-day period but $p \geq 0.05$ in both the nested fortnights. Of these ten instances, two were significant in related variables within the nested fortnights<sup>a</sup>, four were identified in one of the two fortnights directly prior<sup>b</sup>, one was picked up in the fortnight directly after<sup>c</sup>, and three were not found in any fortnight directly before or after<sup>d</sup>.
<sup>a</sup>Ever smoked regularly in the monthly period 11Oct20-07Nov20 ( $p = 0.041$ ). During the fortnights spanning 11Oct20-07Nov20, current smoking status was consistently identified. Impact of long-term health conditions was identified in the 28-days 31Jan21-27Feb21 ( $p = 0.035$ ), where it was marginally significant in the nested fortnight 31Jan21-13Feb21 ( $p = 0.059$ ). Both any long-term health conditions, and disability were flagged as significant in this fortnight.
<sup>b</sup>Any long-term health conditions in 8Nov20-5Dec20; Indoors deprivation index (16Aug20-12Sep20); Sector food production in 31Jan21-27Feb21; Travel abroad (08Nov20-05Dec20; $p = 0.047$ )
<sup>c</sup>Sector finance in 11Oct20-07Nov20
<sup>d</sup>Housing deprivation index in 31Jan21-27Feb21 and 28Mar21-24Apr21 (but this effect did not have an effect after adjusting for overall deprivation index); sector finance in 31Jan21-27Feb21

## Supplementary Figures

**Supplementary Figure 1:**
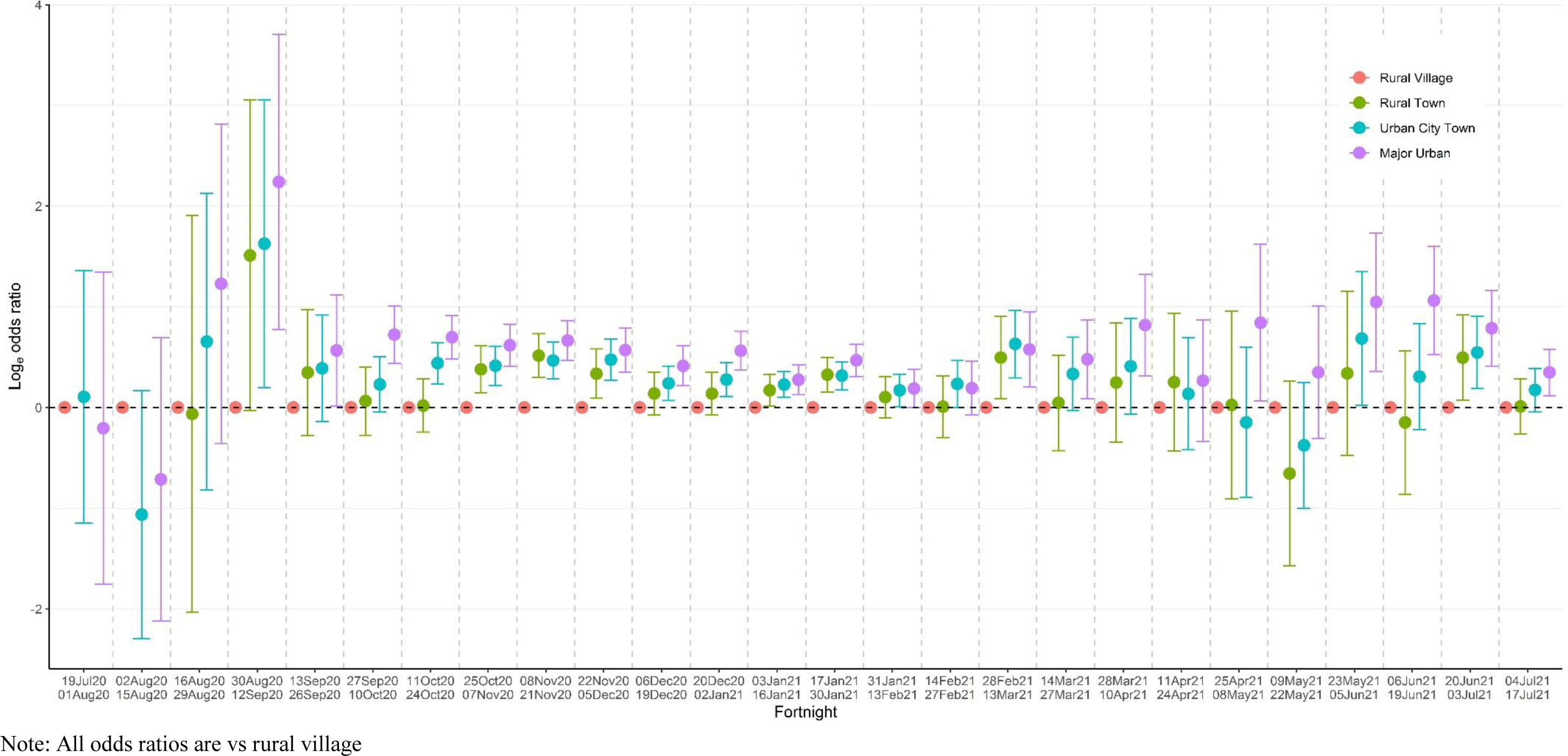
Log odds ratios with 95% confidence intervals for the effect of rural urban classification across the 52 week study period

**Supplementary Figure 2:**
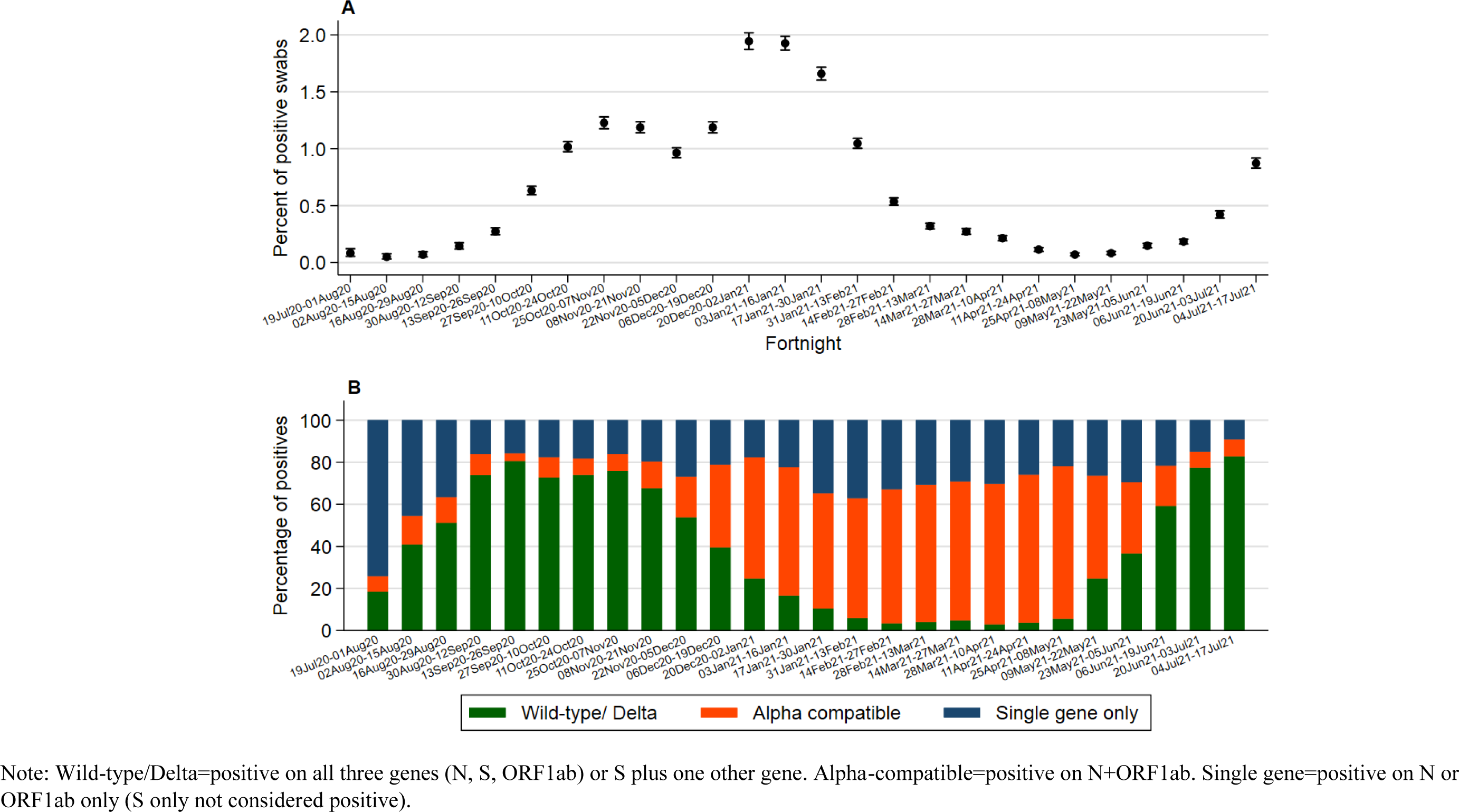
Unadjusted percentage (95% CI) of positive swabs per fortnight (A), and positive swabs split by gene positivity pattern (B)

**Supplementary Figure 3:**
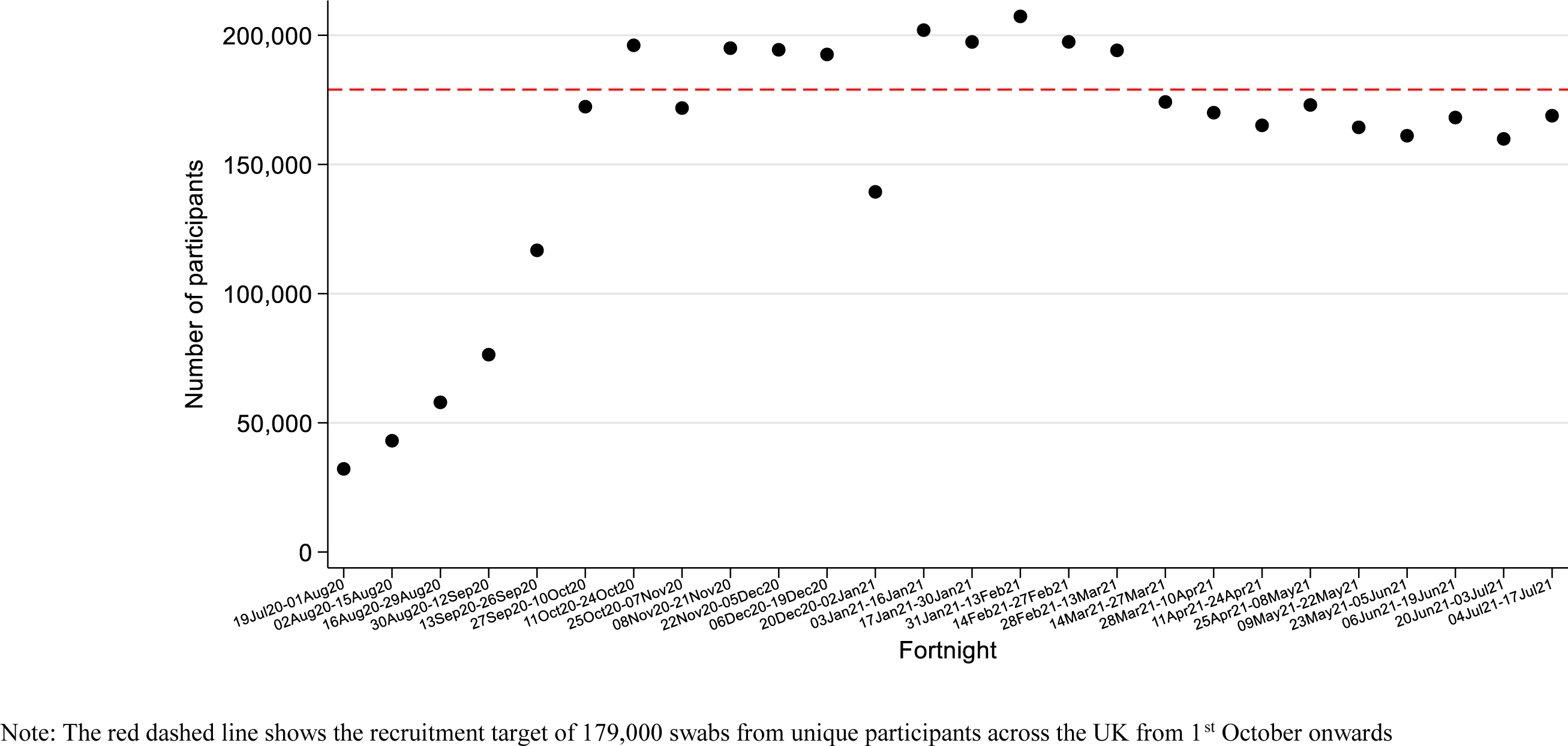
Total number of participants per fortnight

**Supplementary Figure 4:**
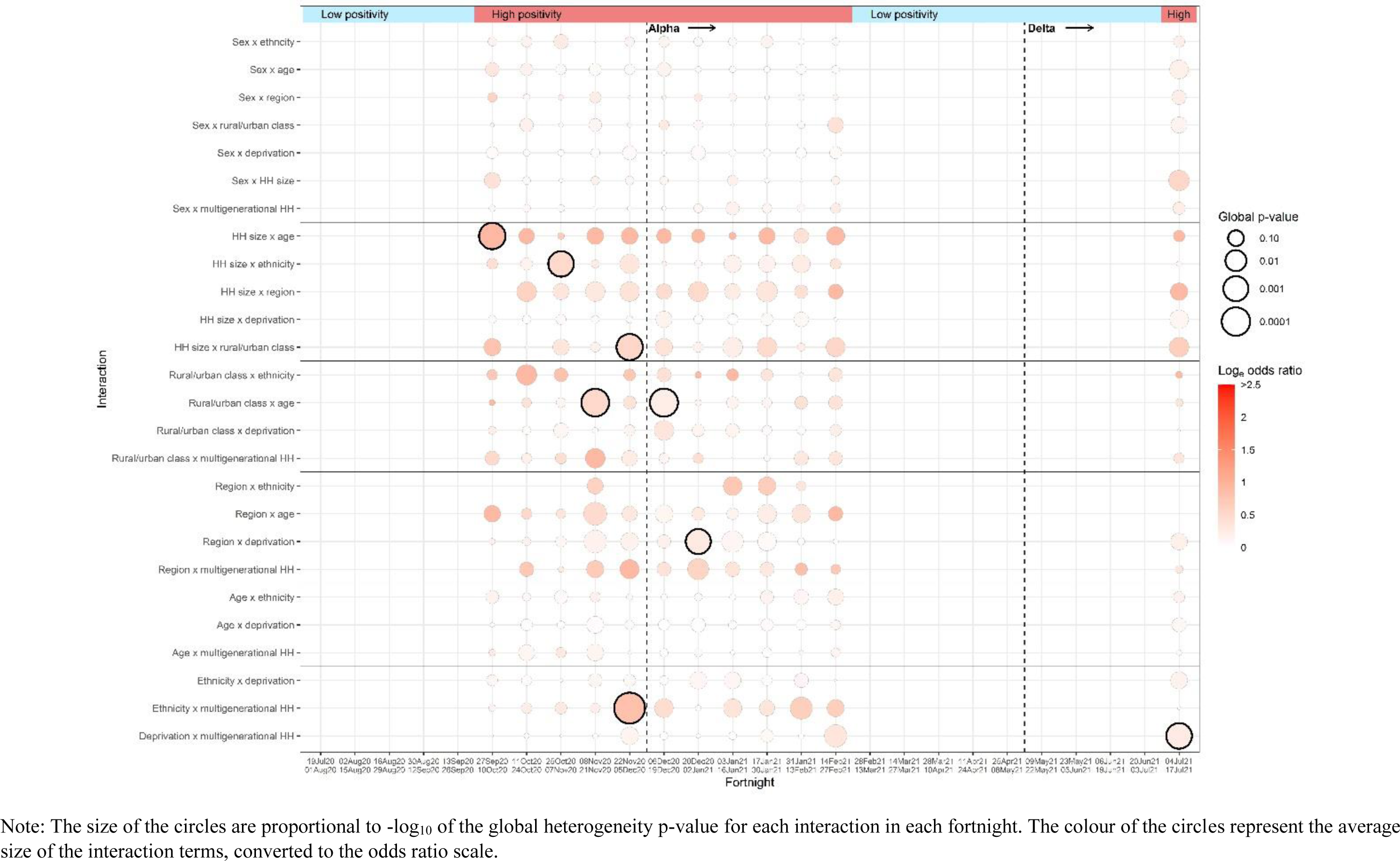
Summary of odds ratio and p-values for interactions between all of the core variables using fortnights.

**Supplementary Figure 5:**
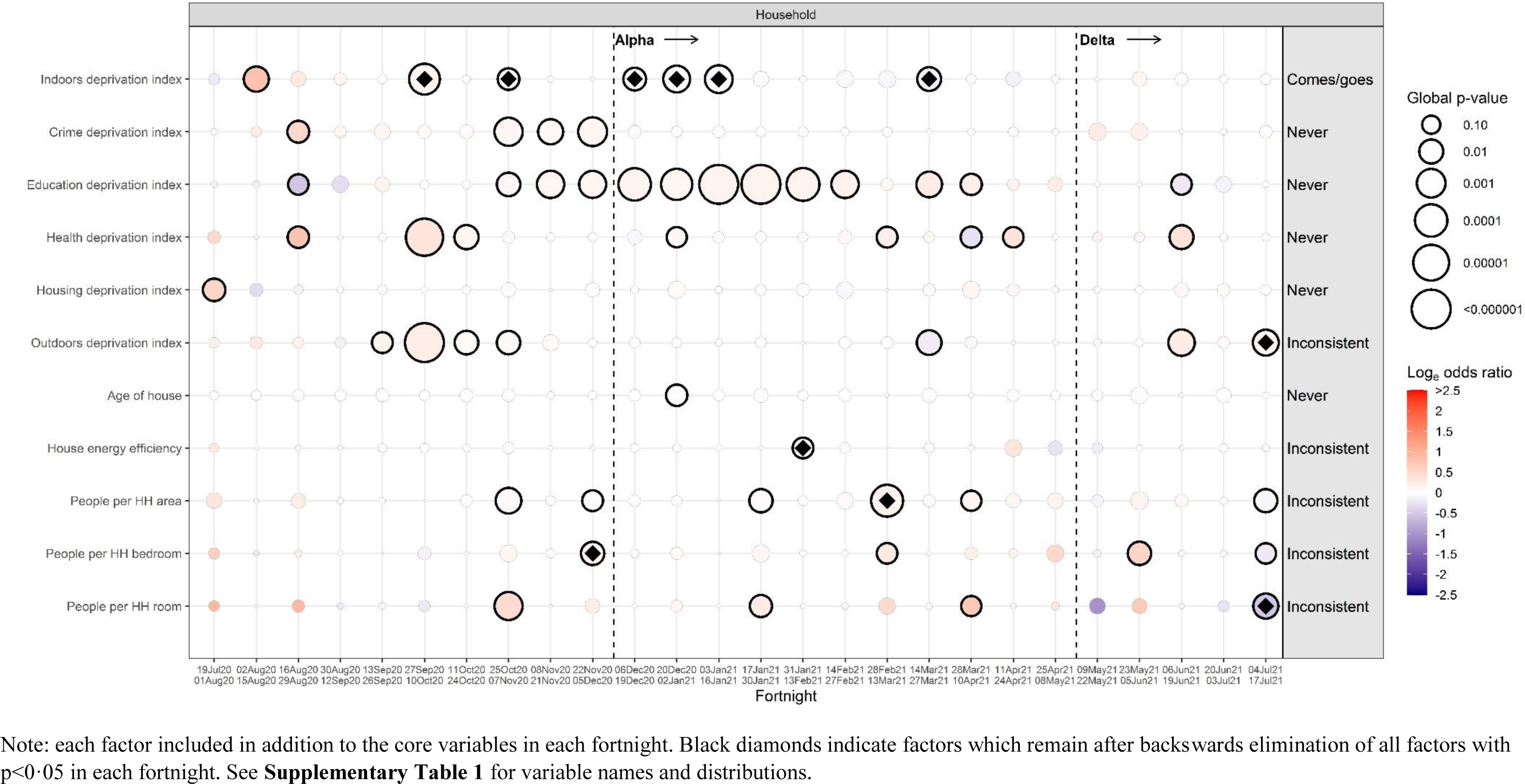
Global hetergeneity p-values per factor from the screening process for household and living enviroment characteristics

**Supplementary Figure 6:**
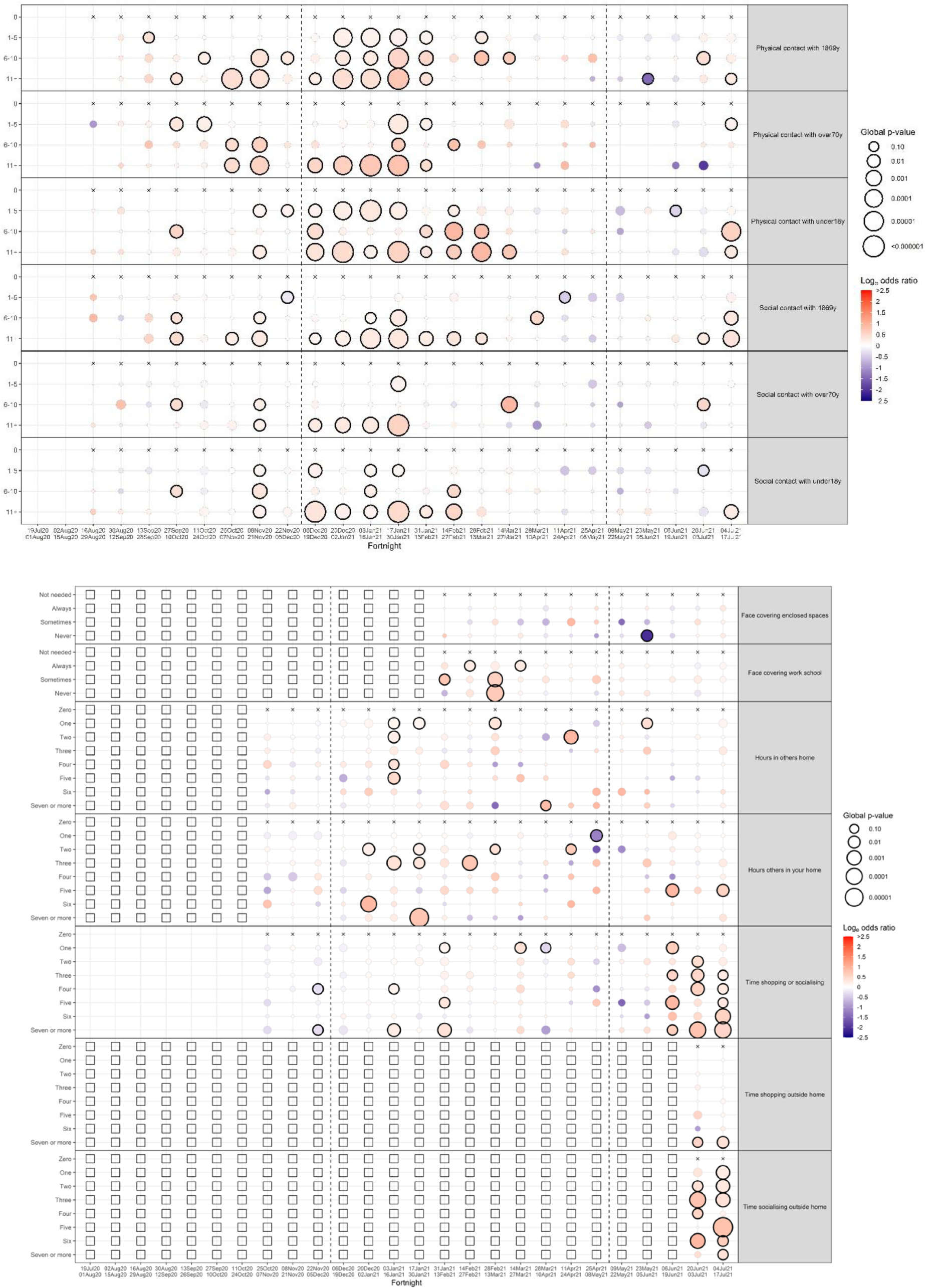
Individual p-values per factor from the screening process for screening characteristics

**Supplementary Figure 7A:**
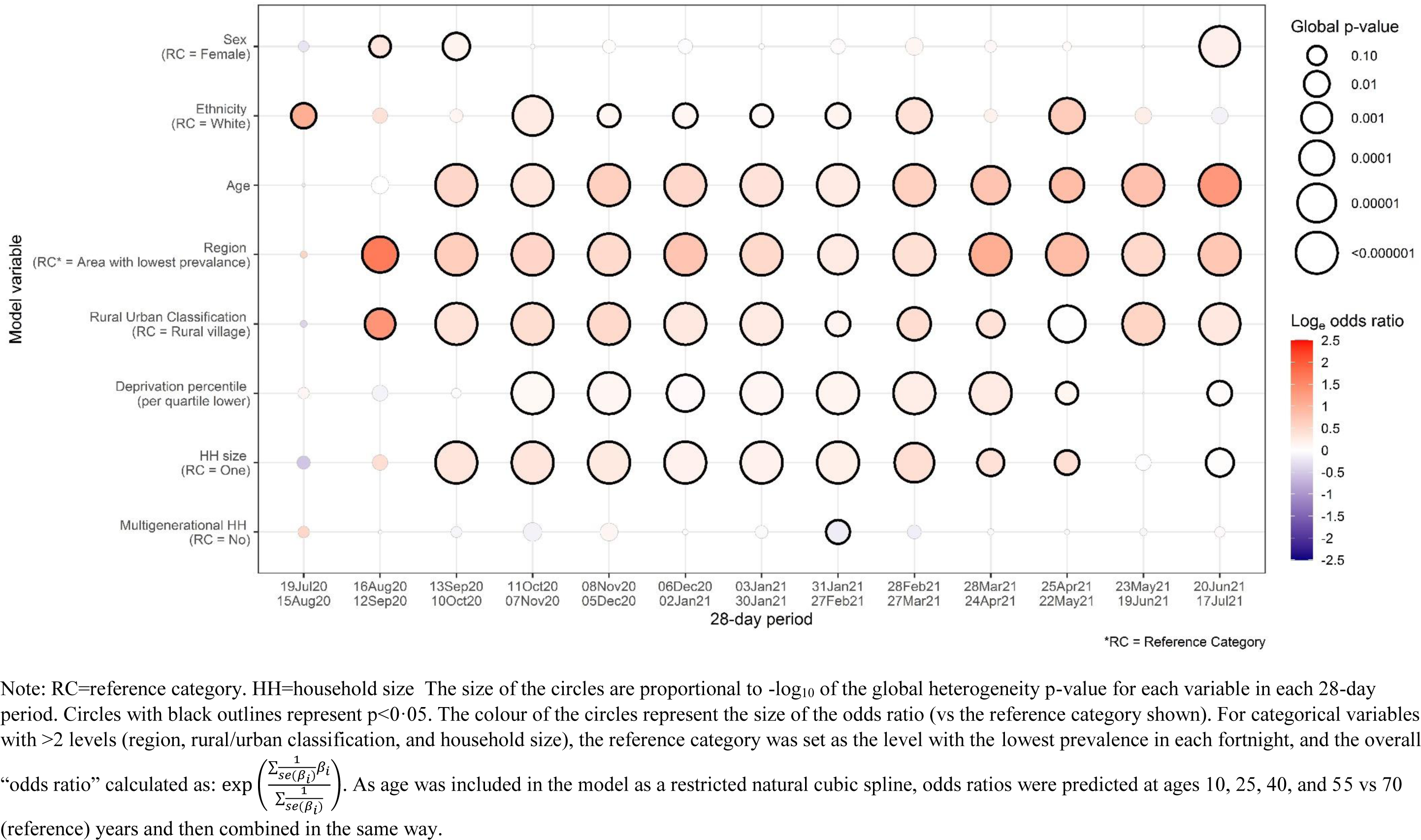
Summary of odds ratios and p-values for the 8 core variables over 28 day periods

**Supplementary Figure 7B:**
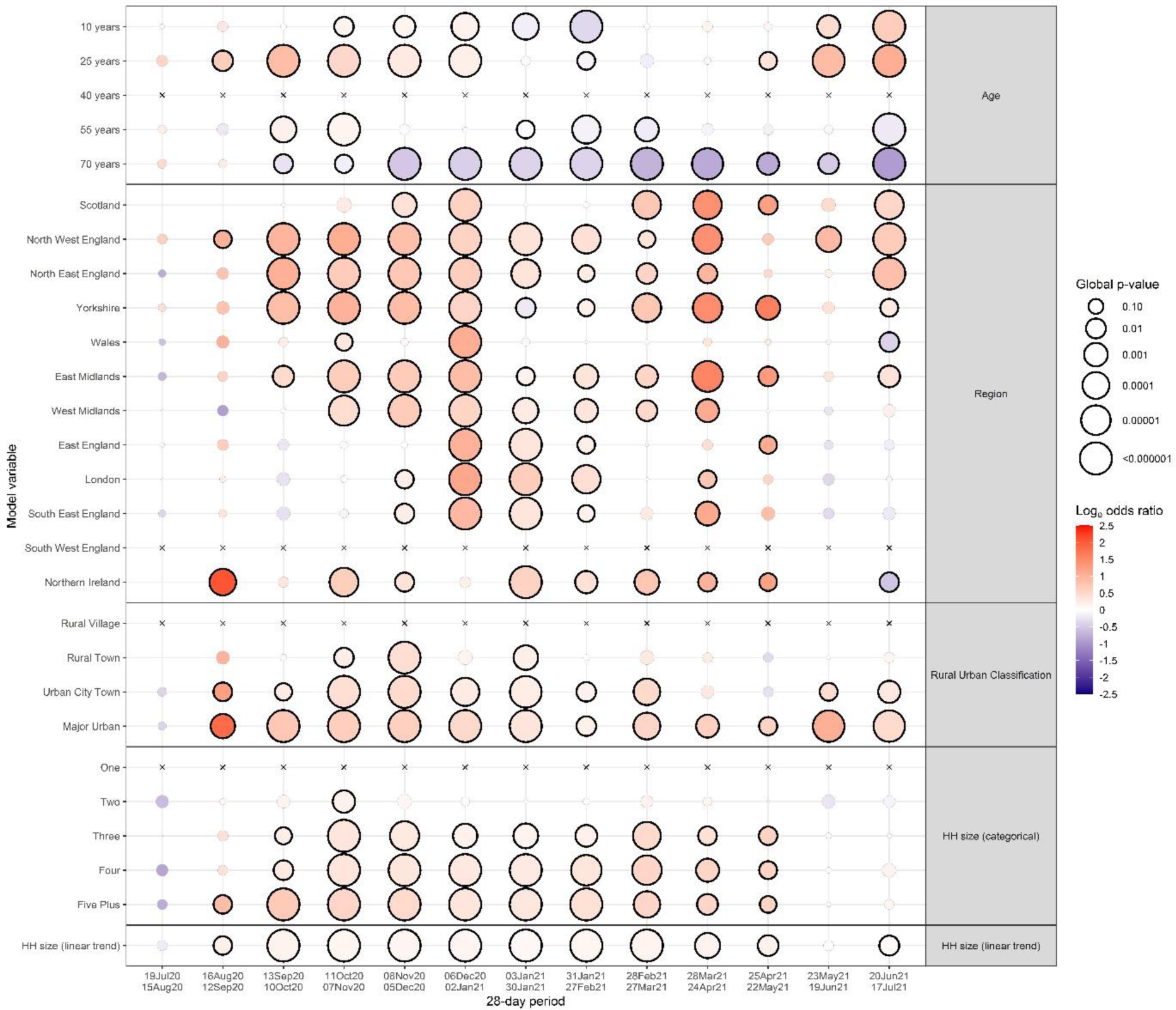
Summary of odds ratios and p-values for the individual levels of the 8 core variables over 28 day periods

**Supplementary Figure 8:**
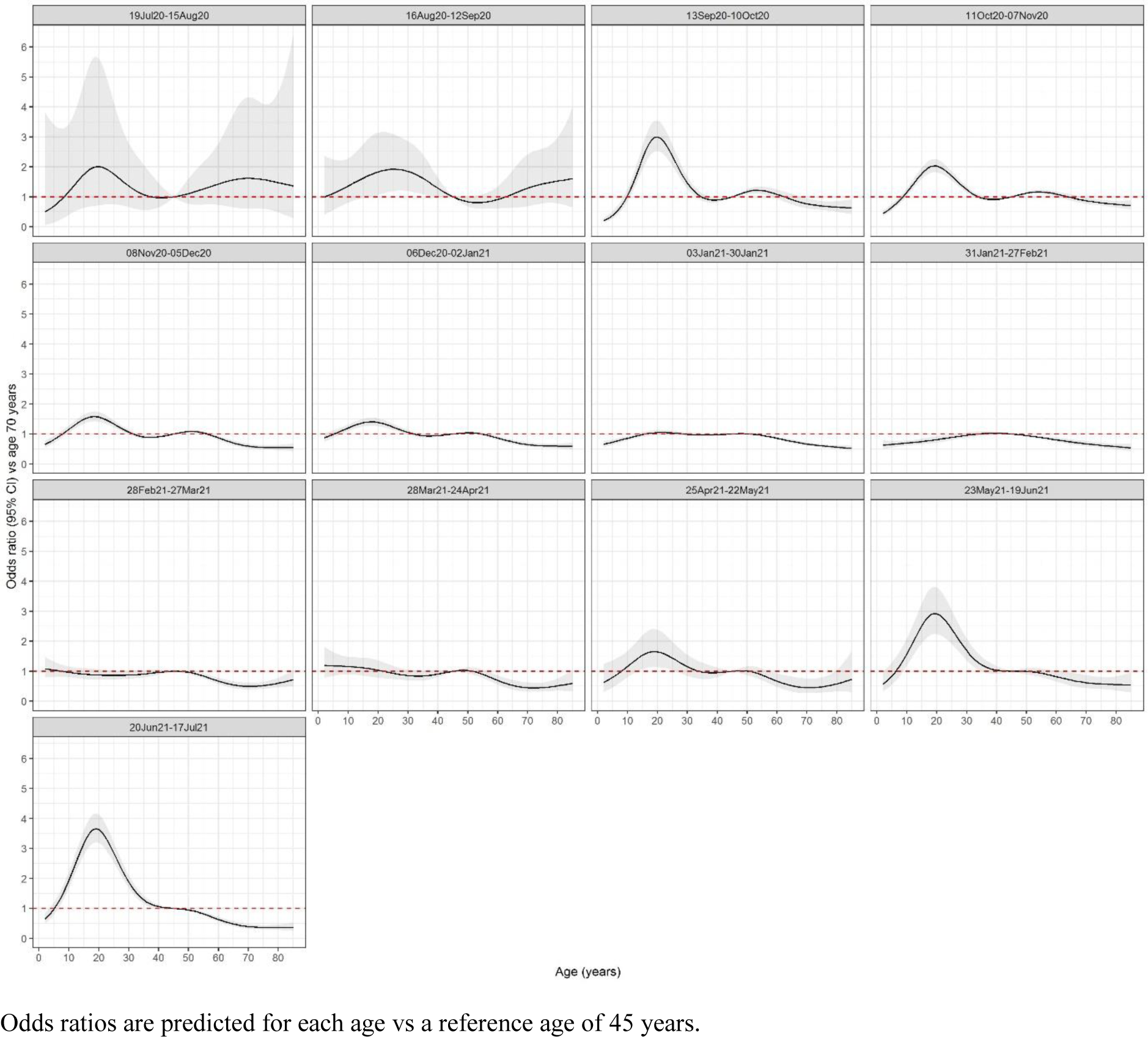
Adjusted effect of age (years) on positivity using 28-day periods.

**Supplementary Figure 9A:**
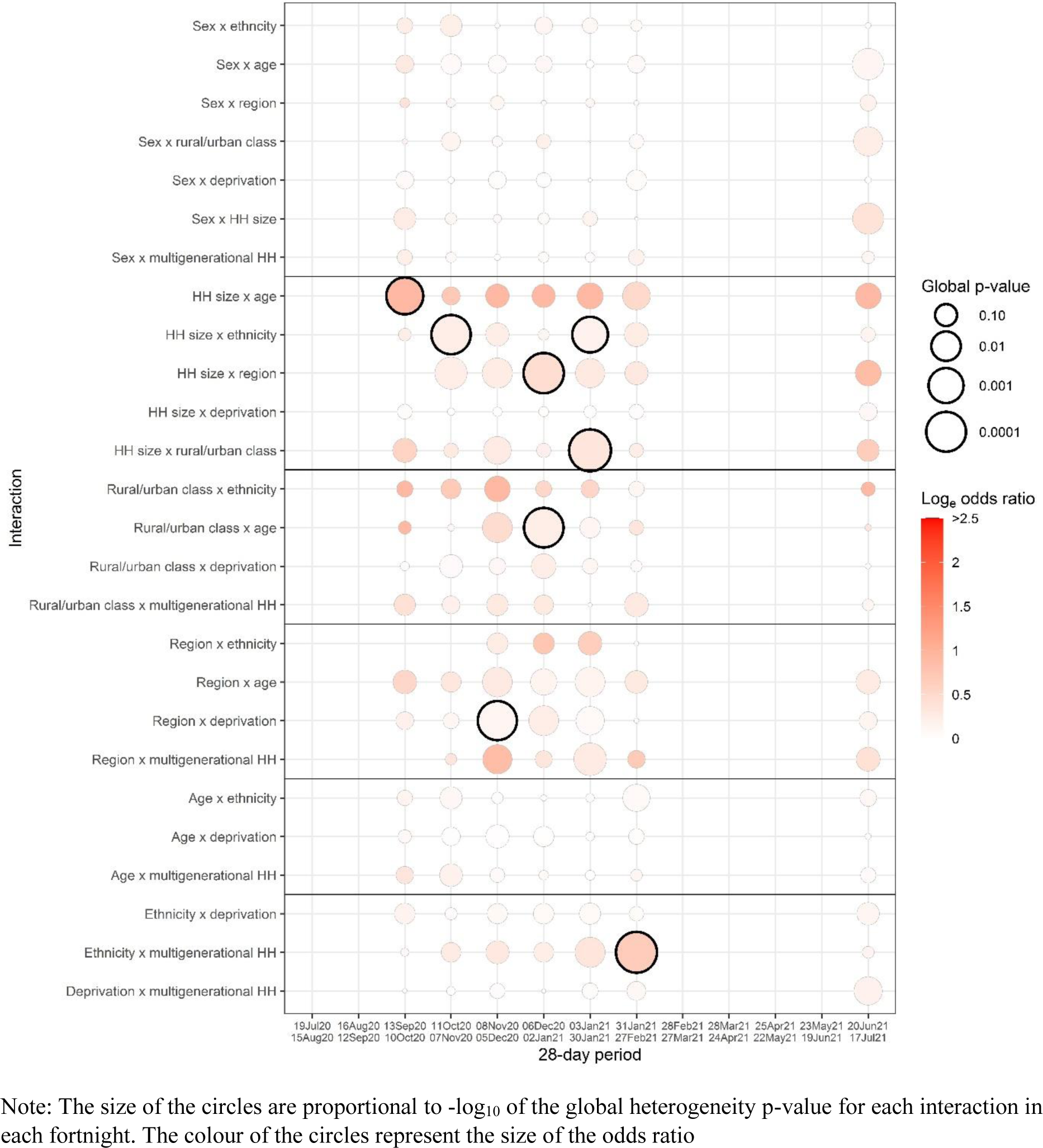
Summary of odds ratio and p-values for interactions between all of the core variables for 28 day periods.

**Figure 9B:**
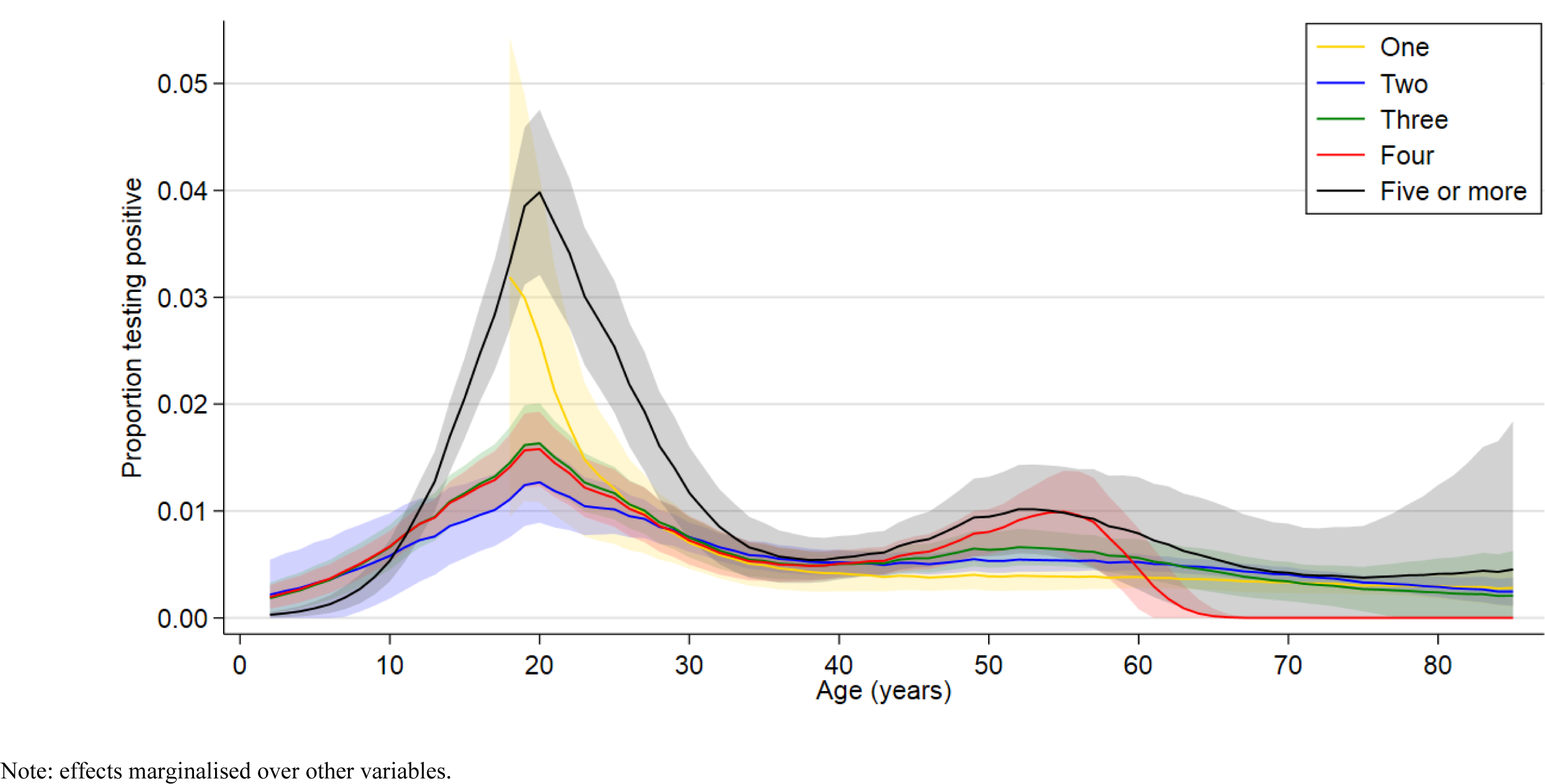
Effect of interaction of age by household size in the 28-day period 13 September to 10^th^ October

**Figure 9C:**
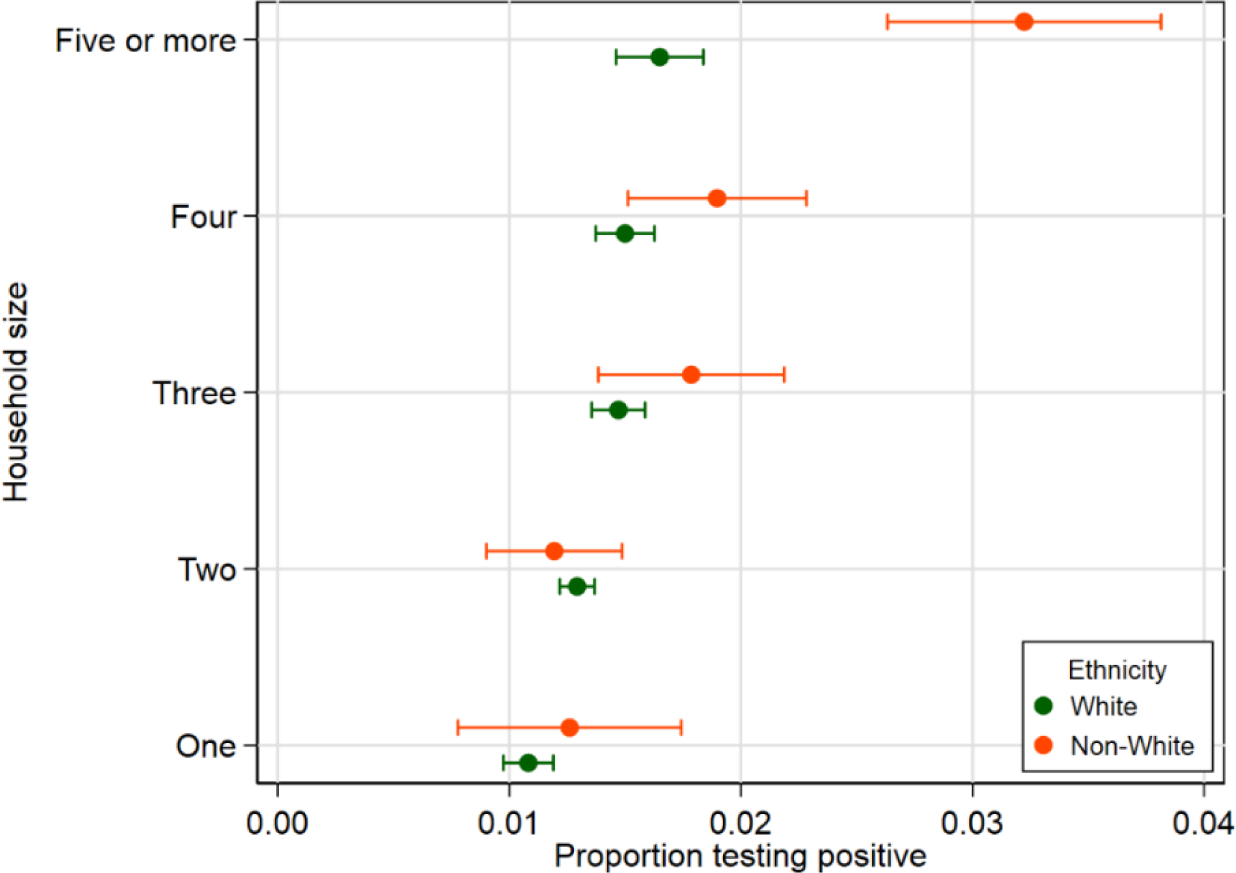
Effect of interaction of ethnicity by household size in the 28-day period 11^th^ October 2020 to 7^th^ November 2020

**Figure 9D:**
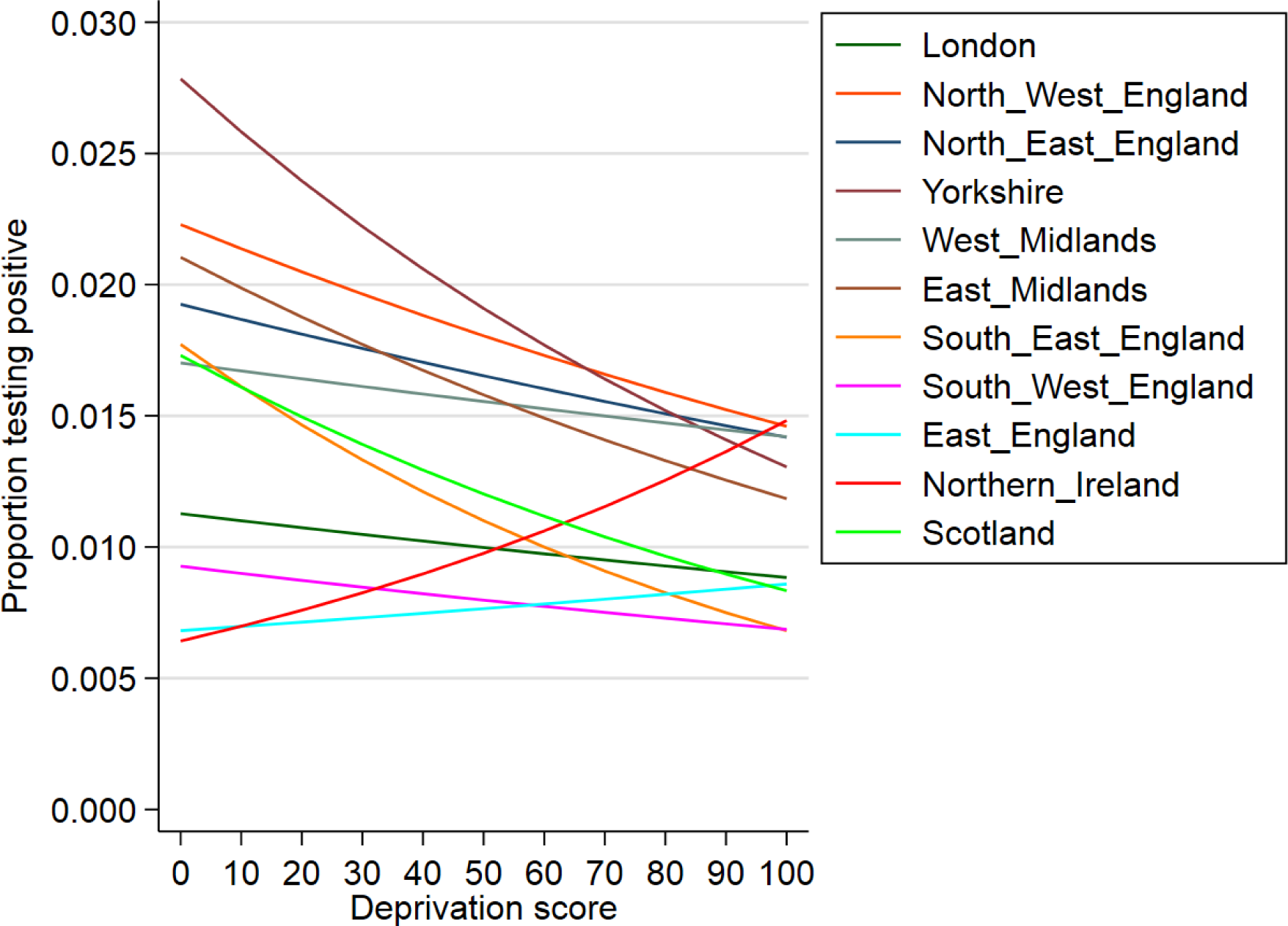
Effect of interaction of region by deprivation score in the 28-day period 8^th^ November 2020 to 5^th^ December 2020

**Figure 9E:**
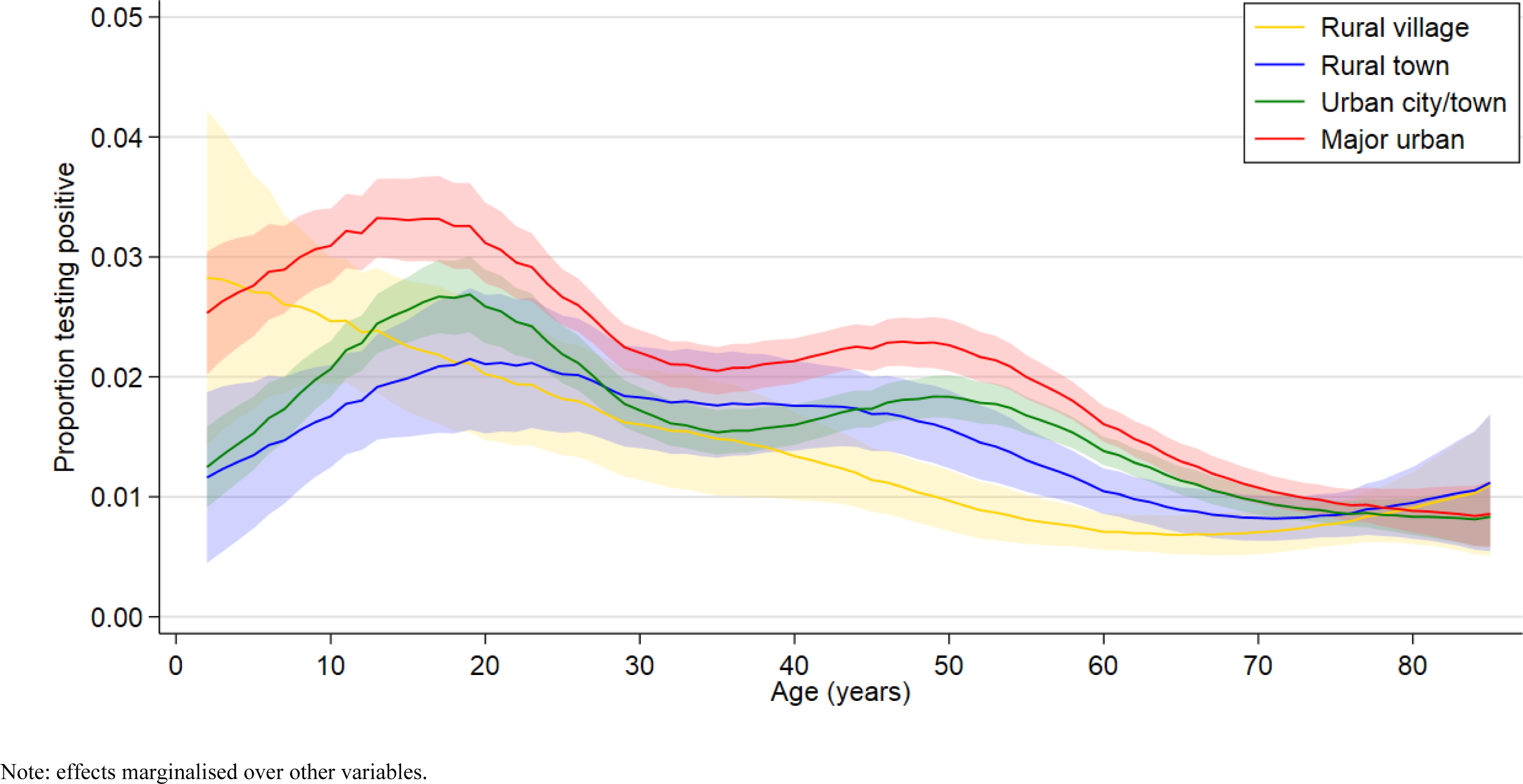
Effect of interaction of rural urban classification by age in the 28-day period 6^th^ December 2020 to 2^nd^ January 2021

**Figure 9F:**
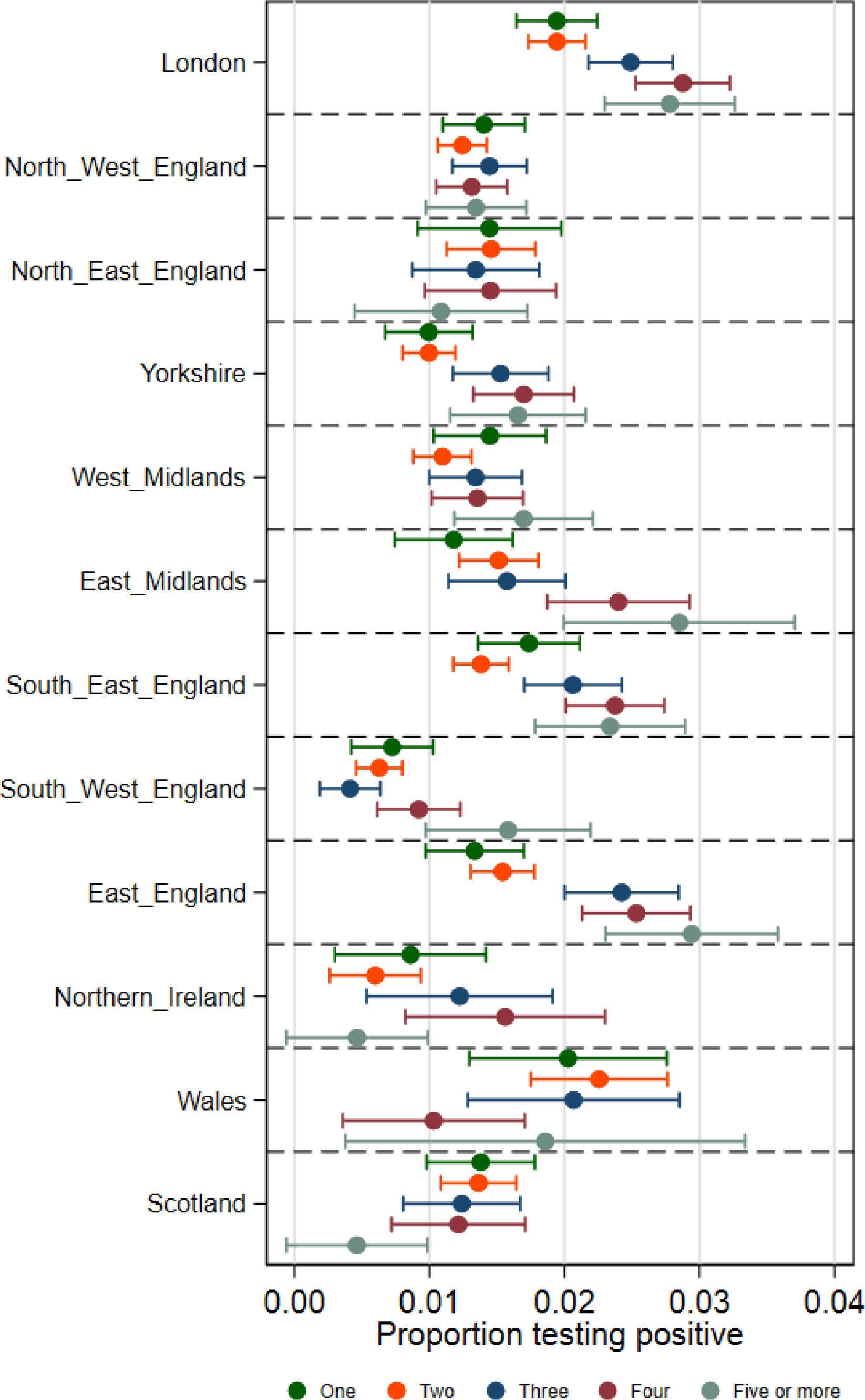
Effect of interaction of region by household size in the 28-day period 6^th^ December 2020 to 2^nd^ January 2021

**Figure 9G:**
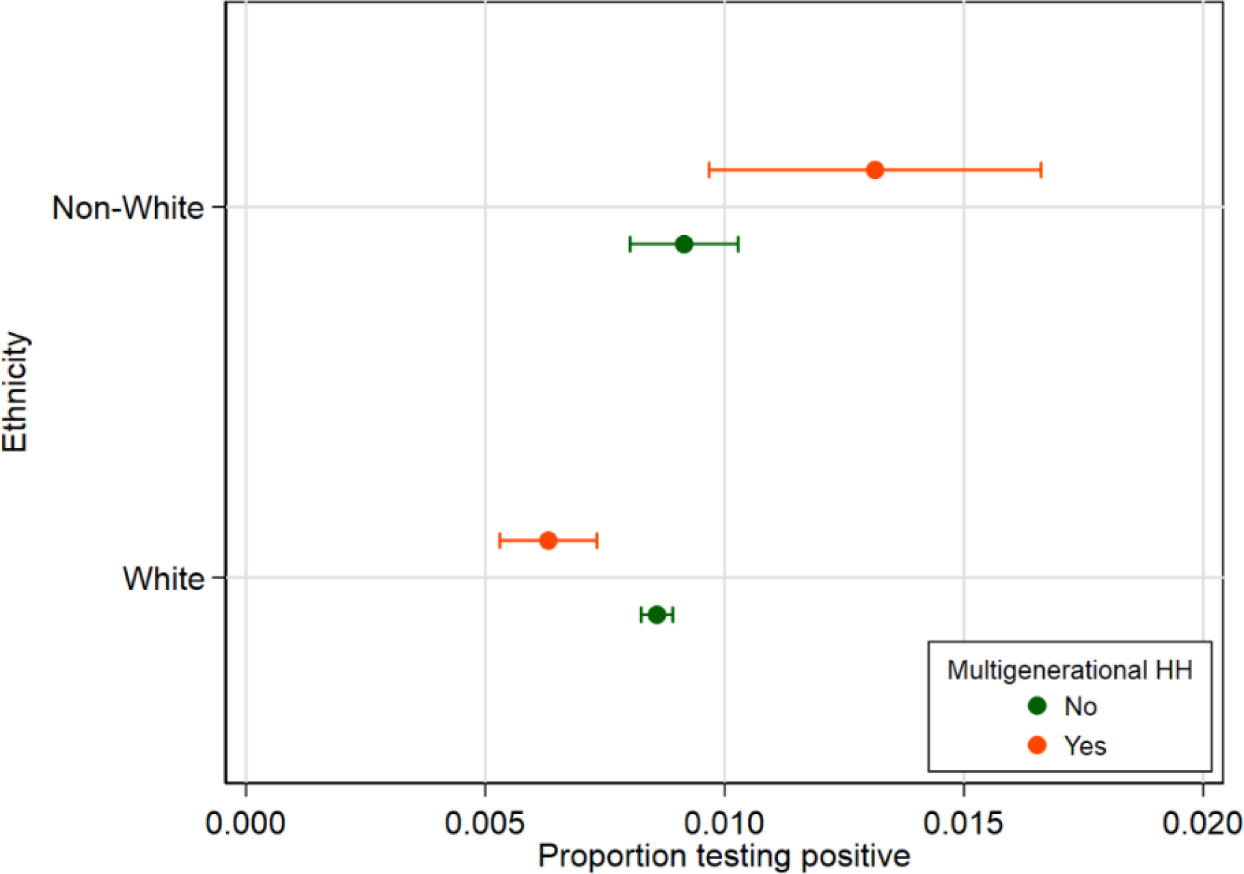
Effect of interaction of ethnicity by multigenerational households in the 28-day period 31^st^ January 2021 to 27^th^ February 2021

**Supplementary Figure 10A:**
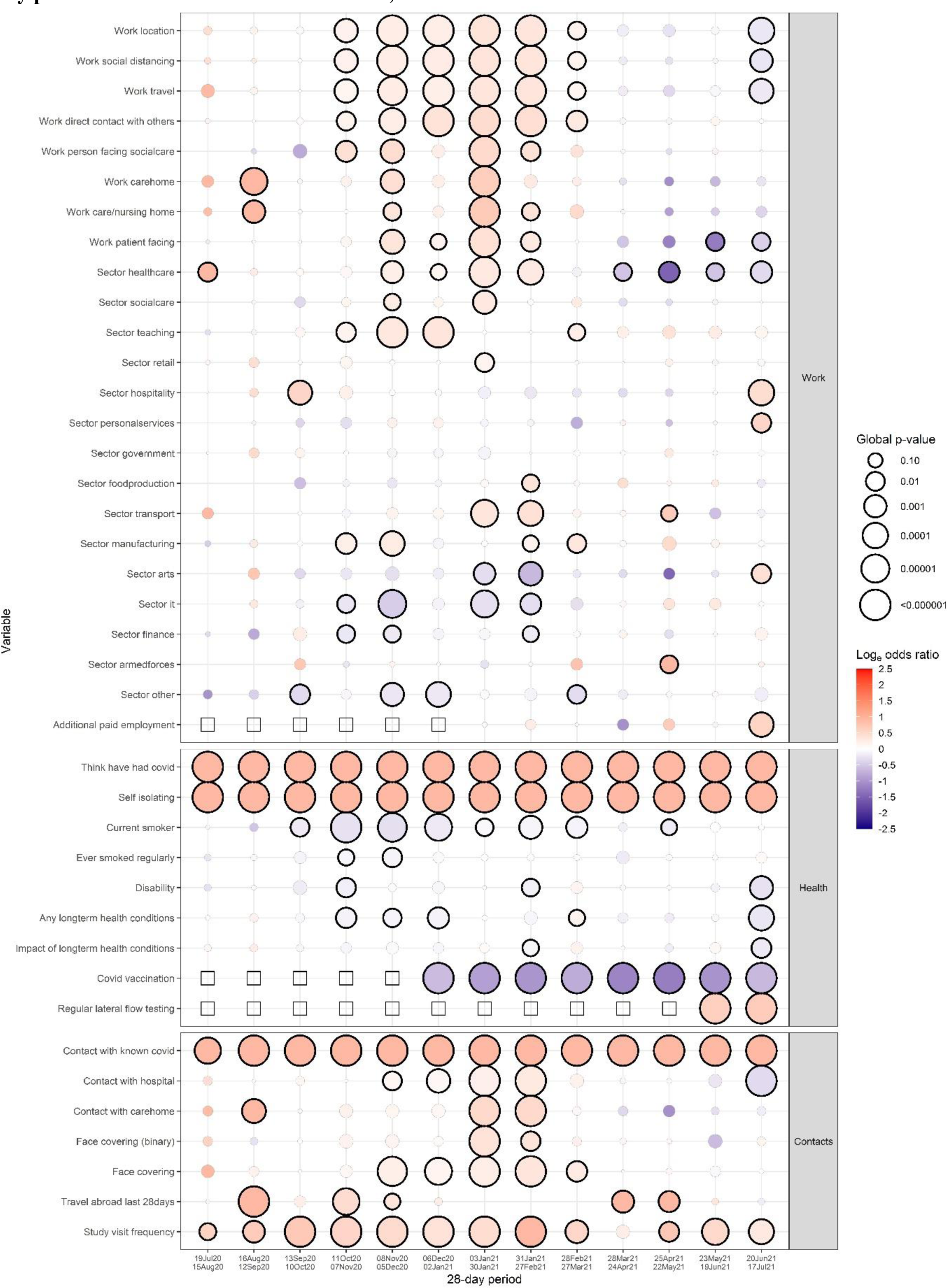
Global heterogeneity p-values per factor from the screening process for 28- day periods for characetrics based on work, health status and contacts

**Supplementary Figure 10B:**
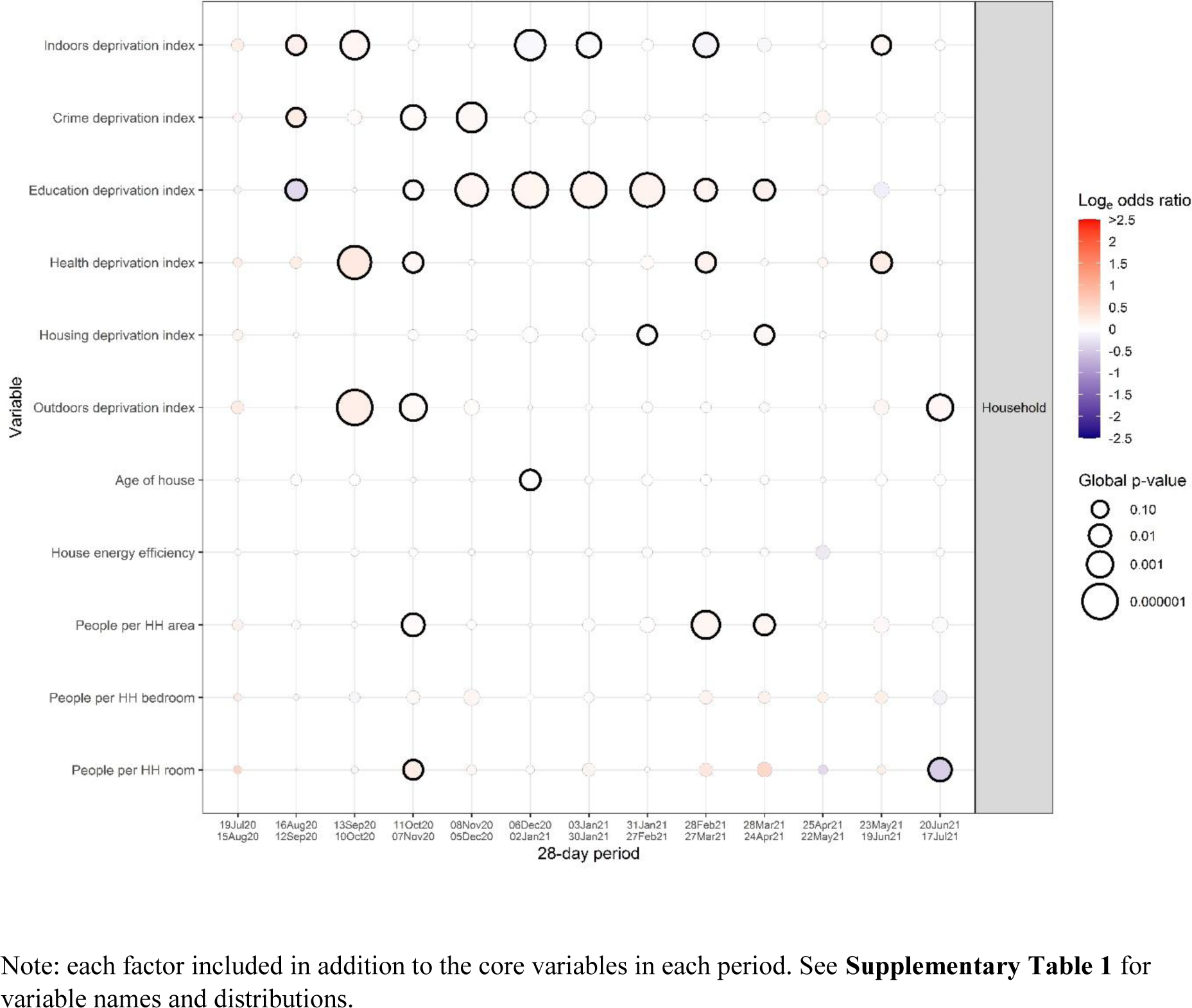
Global heterogeneity p-values per factor from the screening process for 28- day periods for characteristics based on household and living environment

**Supplementary Figure 11:**
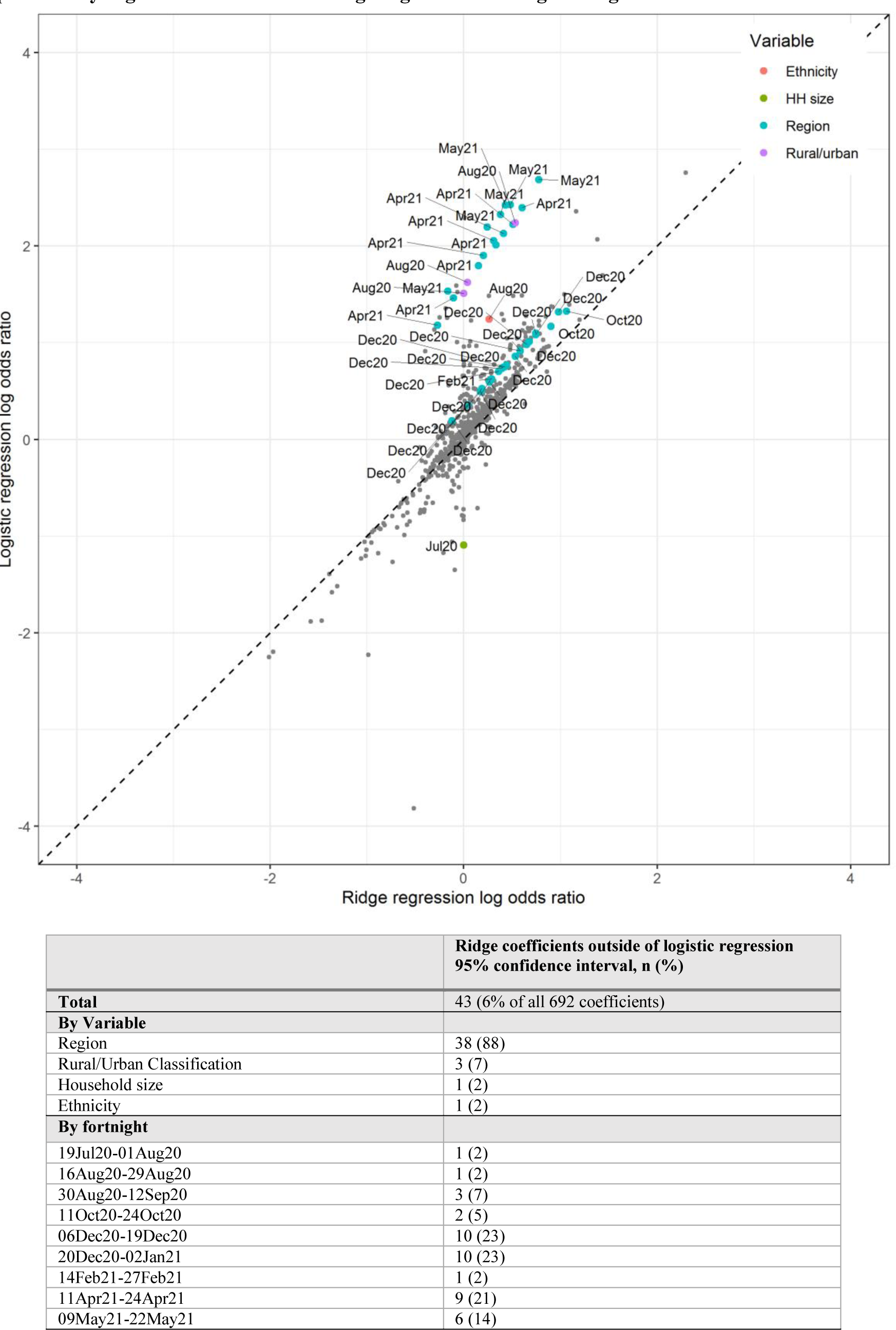
Results from ridge regression and logistic regression

**Supplementary Figure 12:**
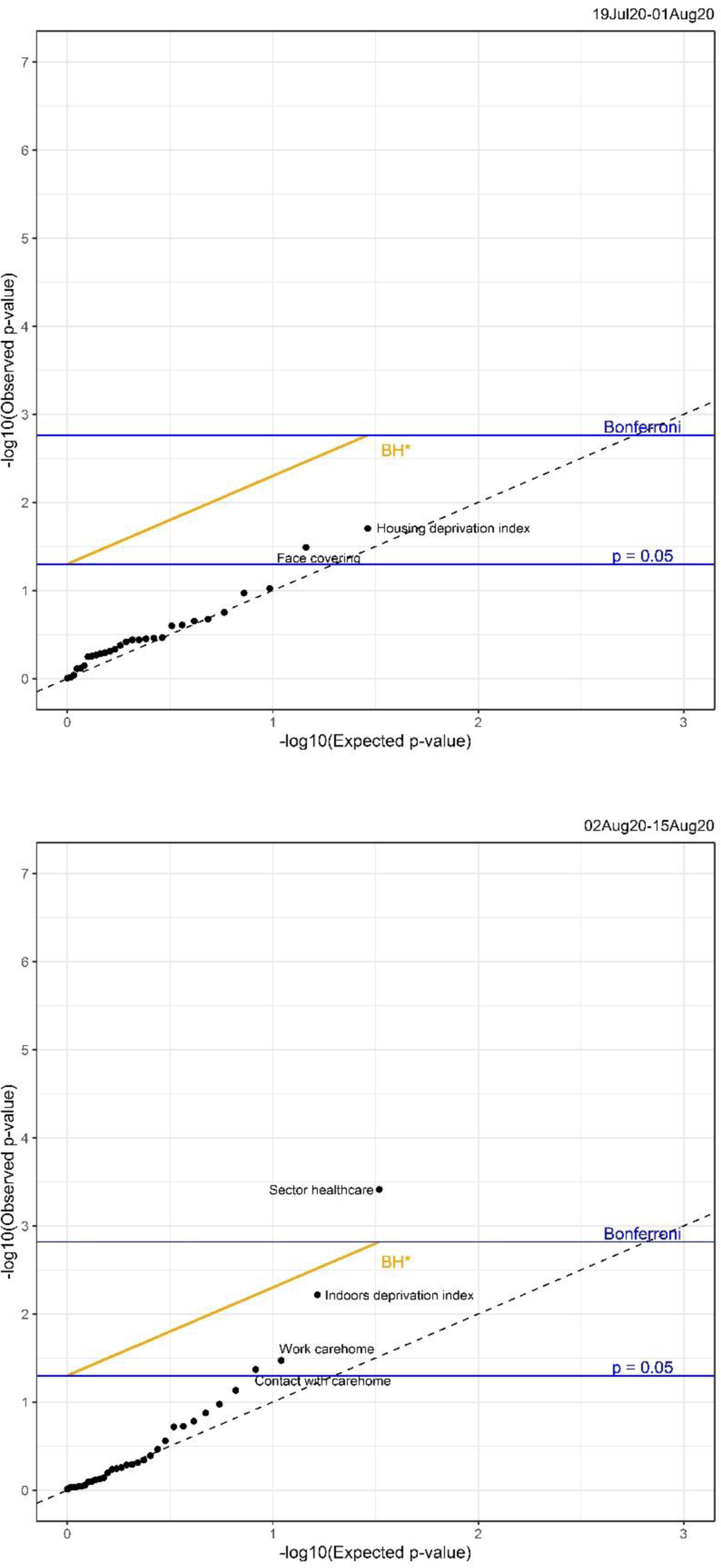
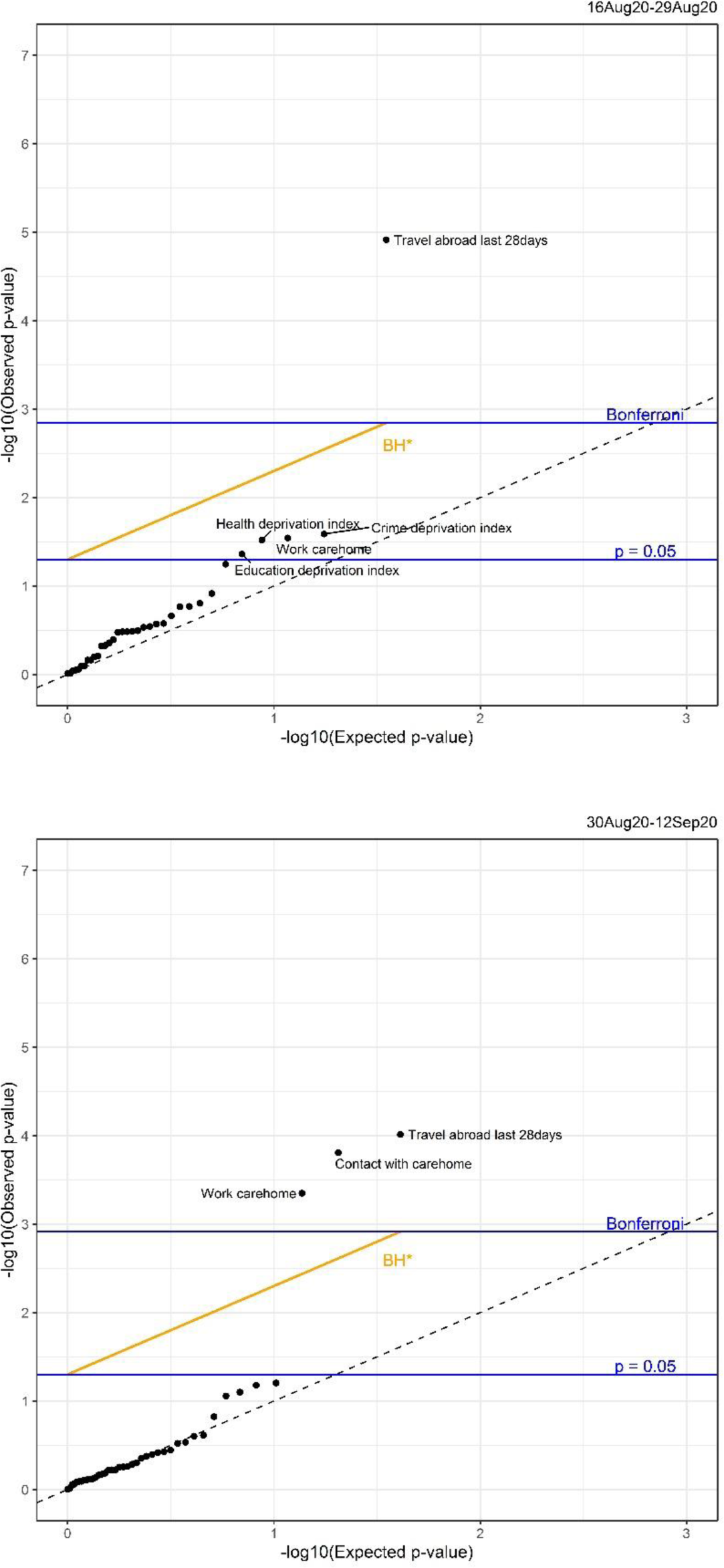
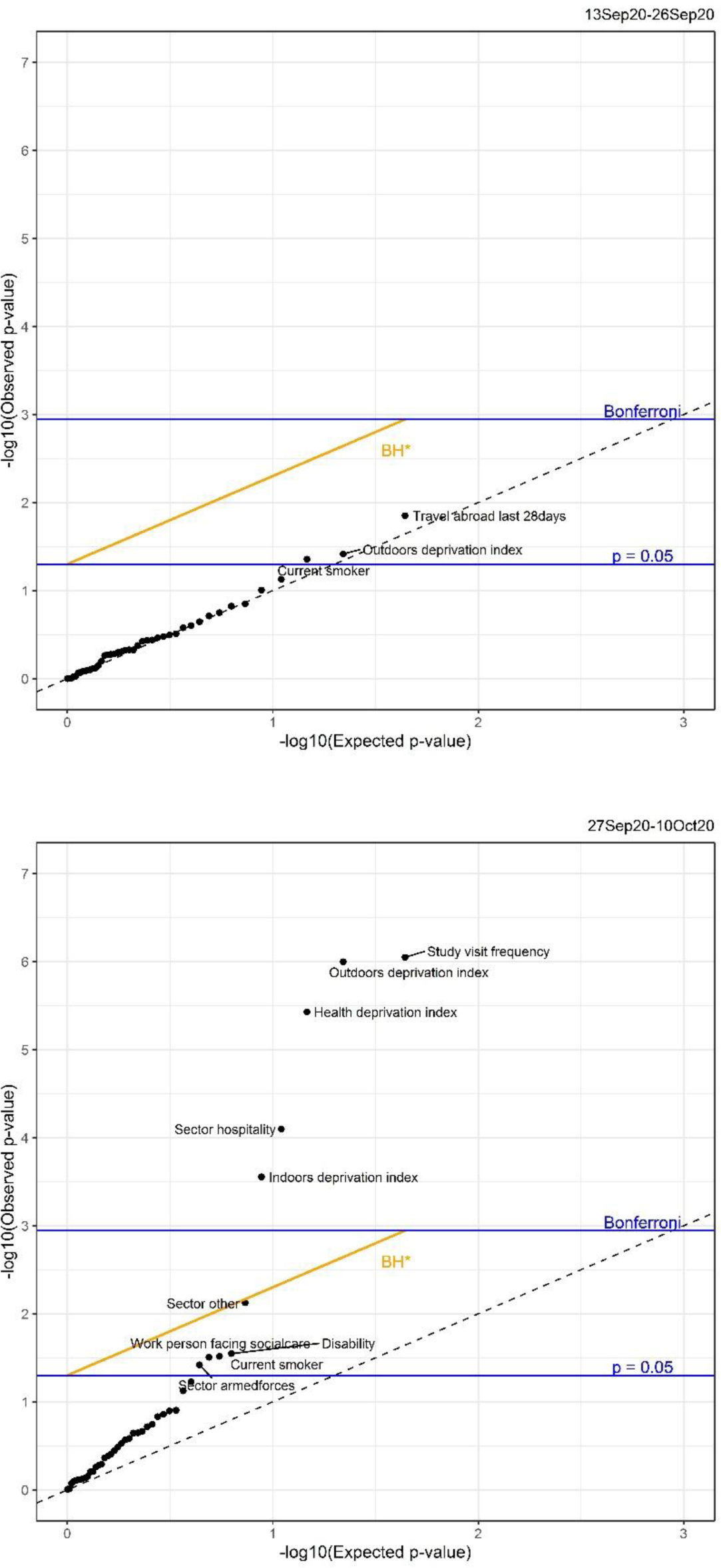
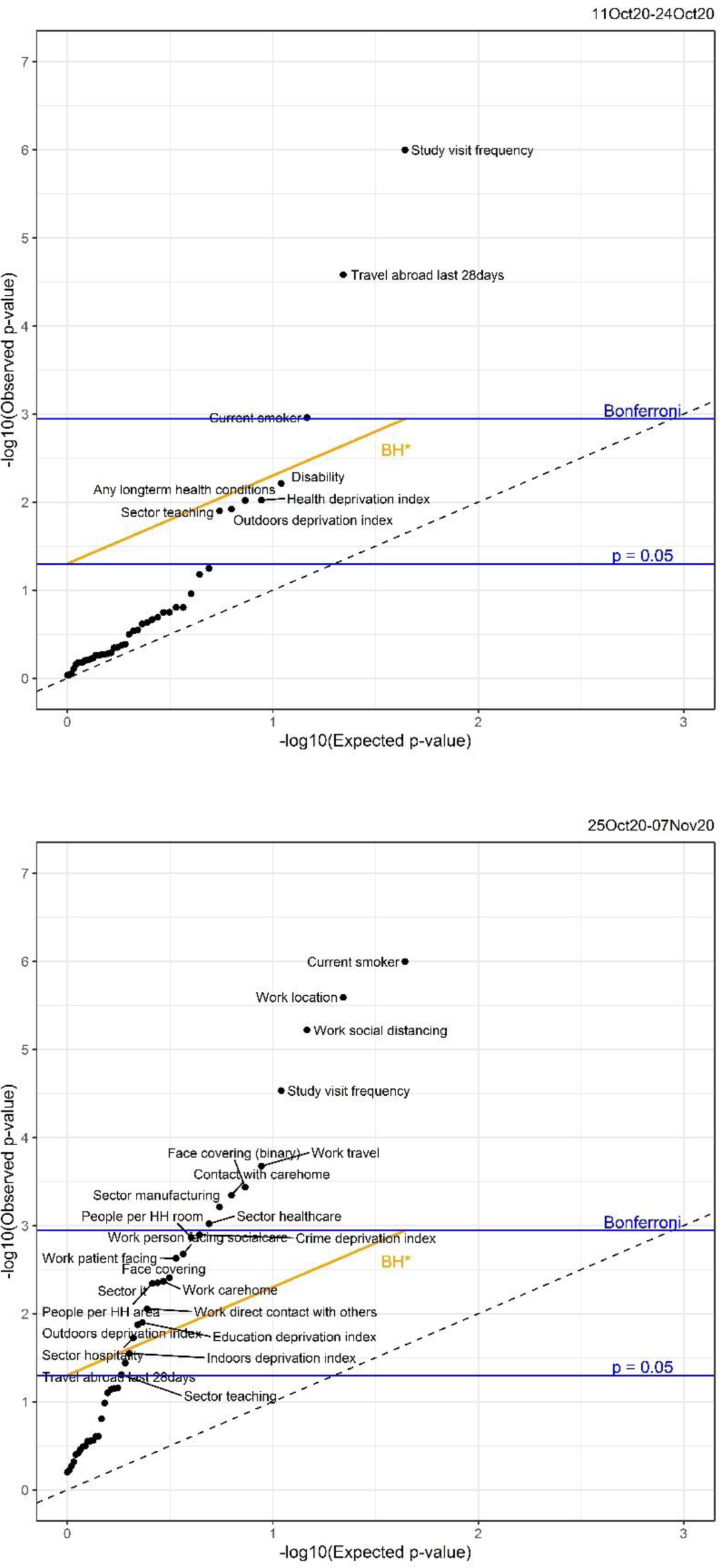
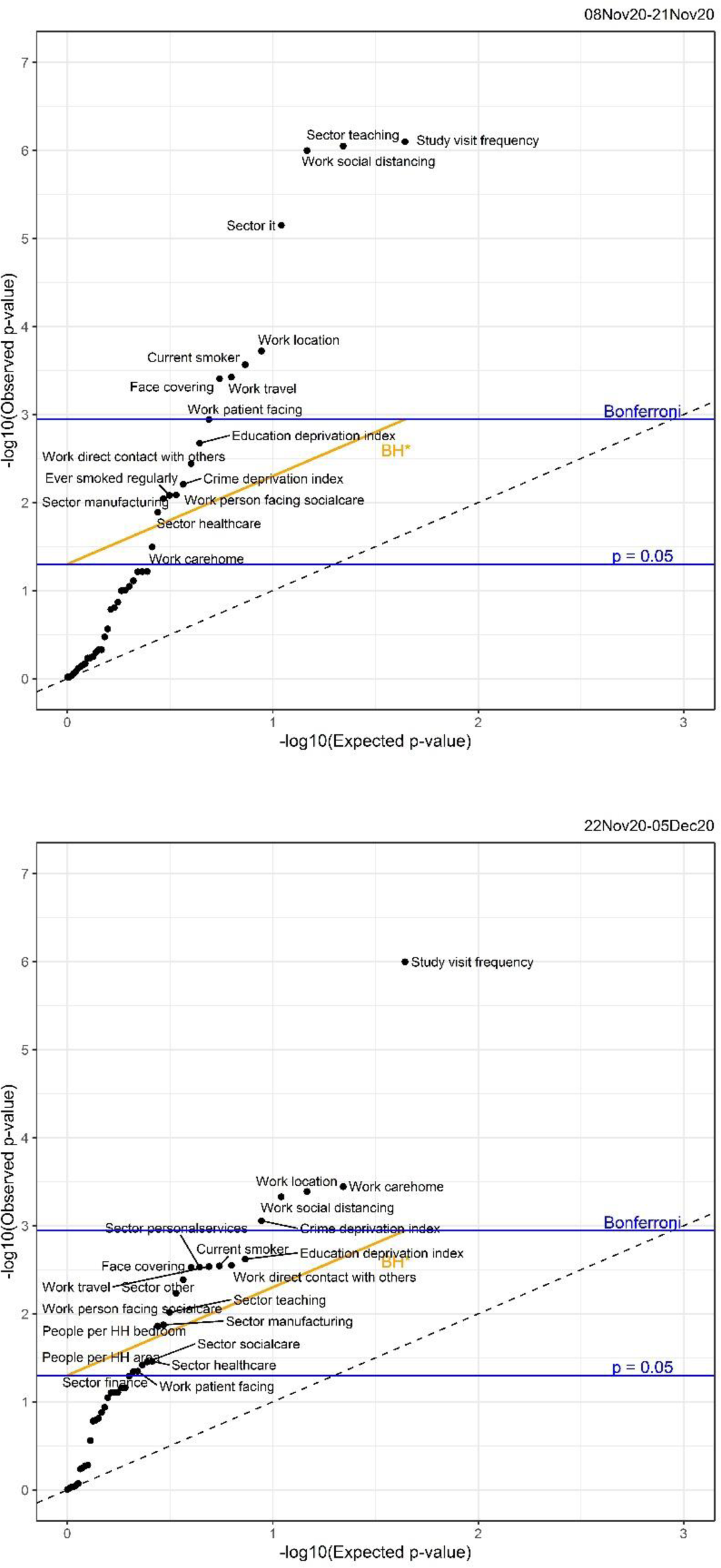
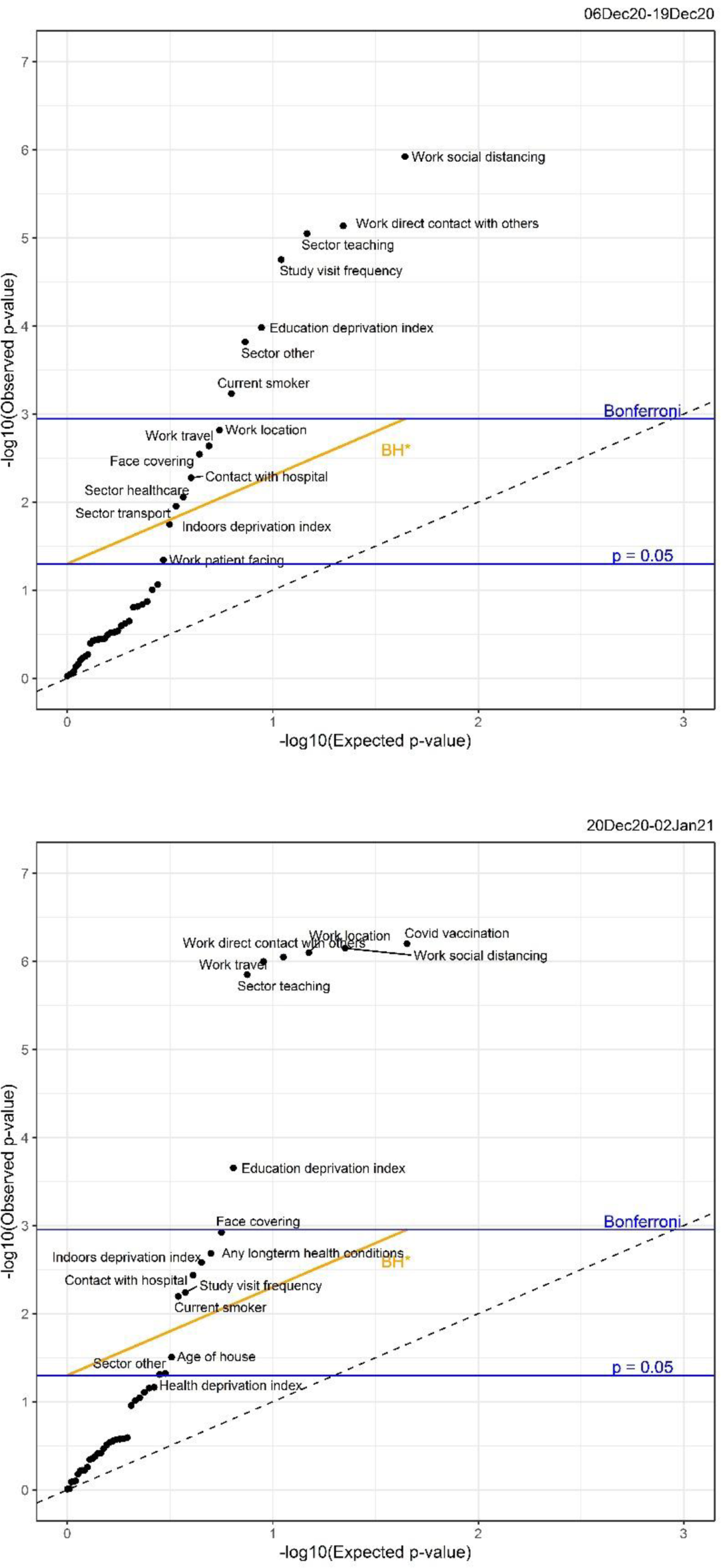
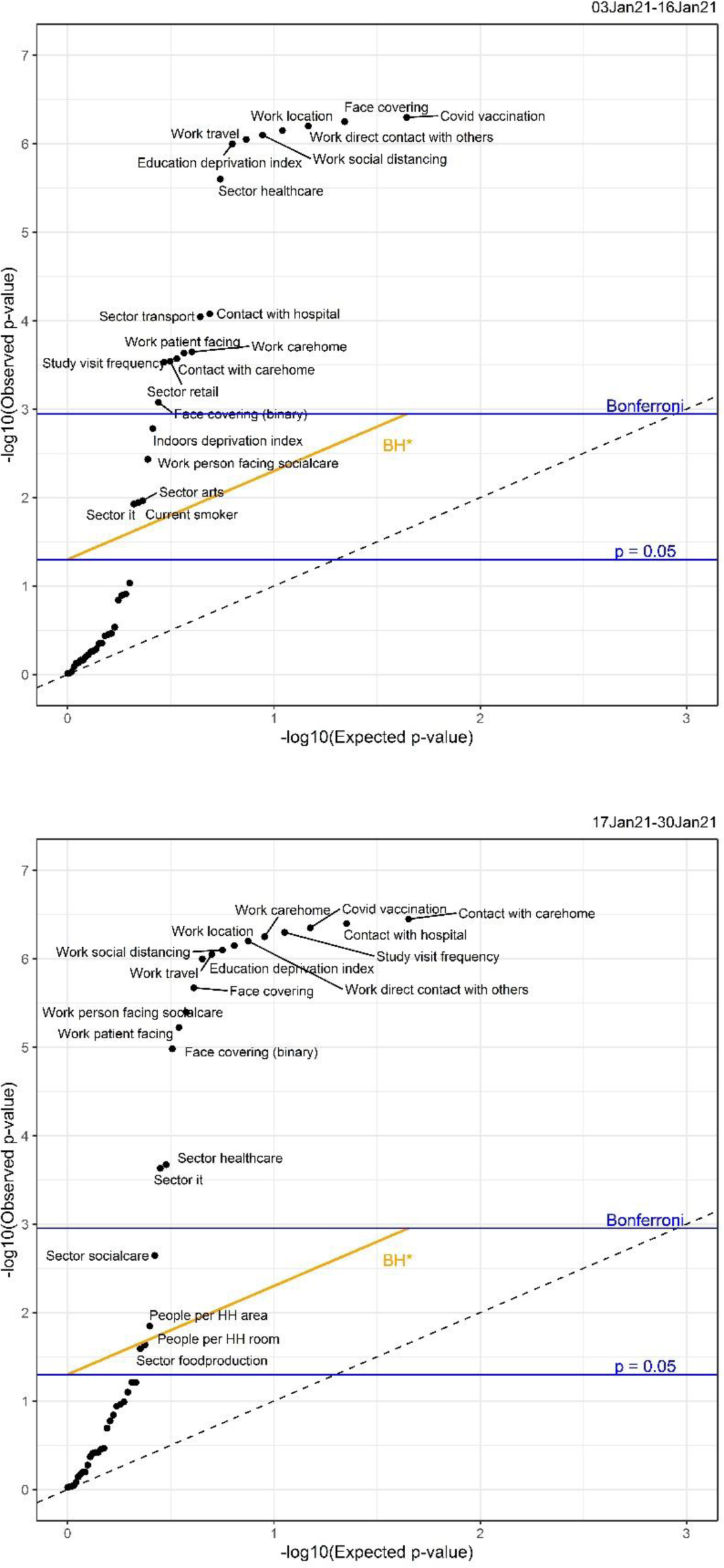
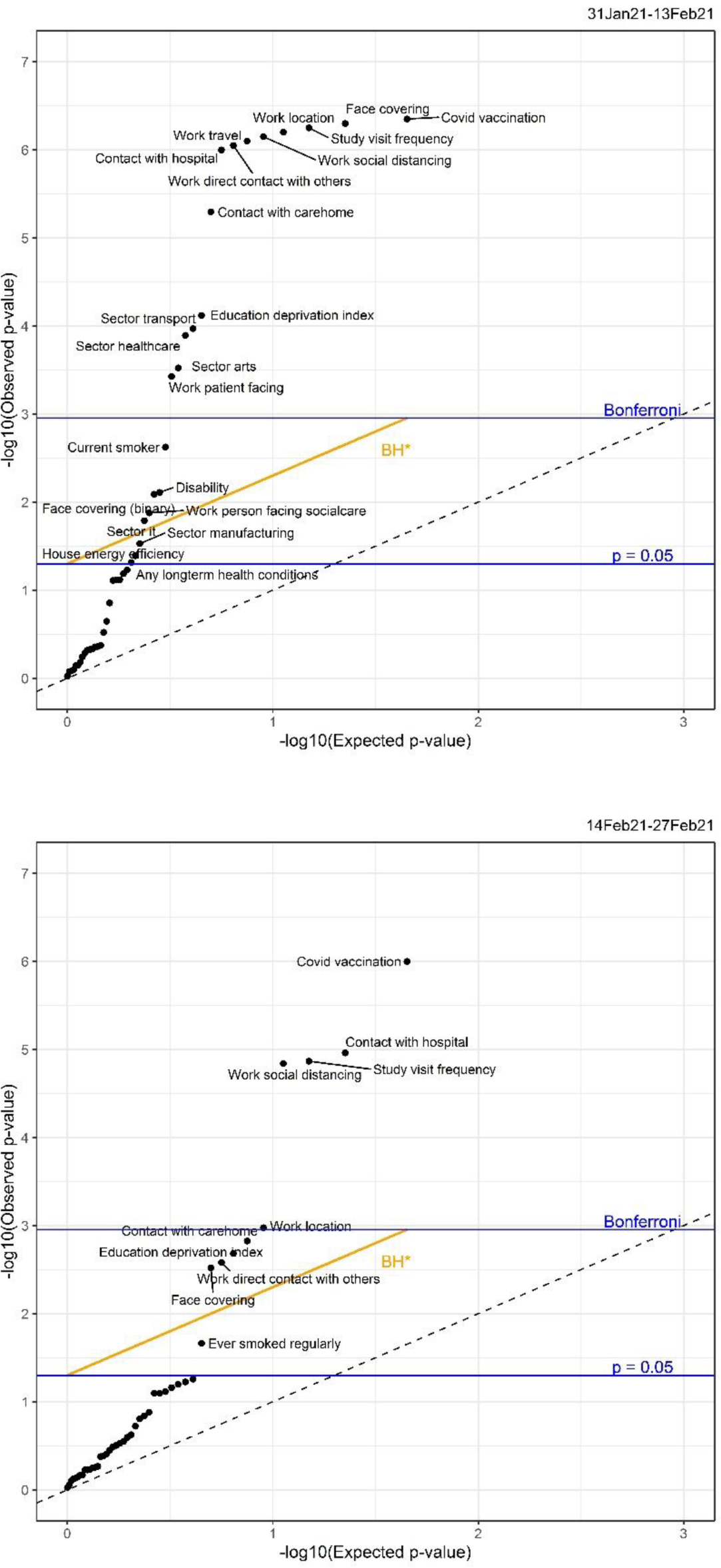
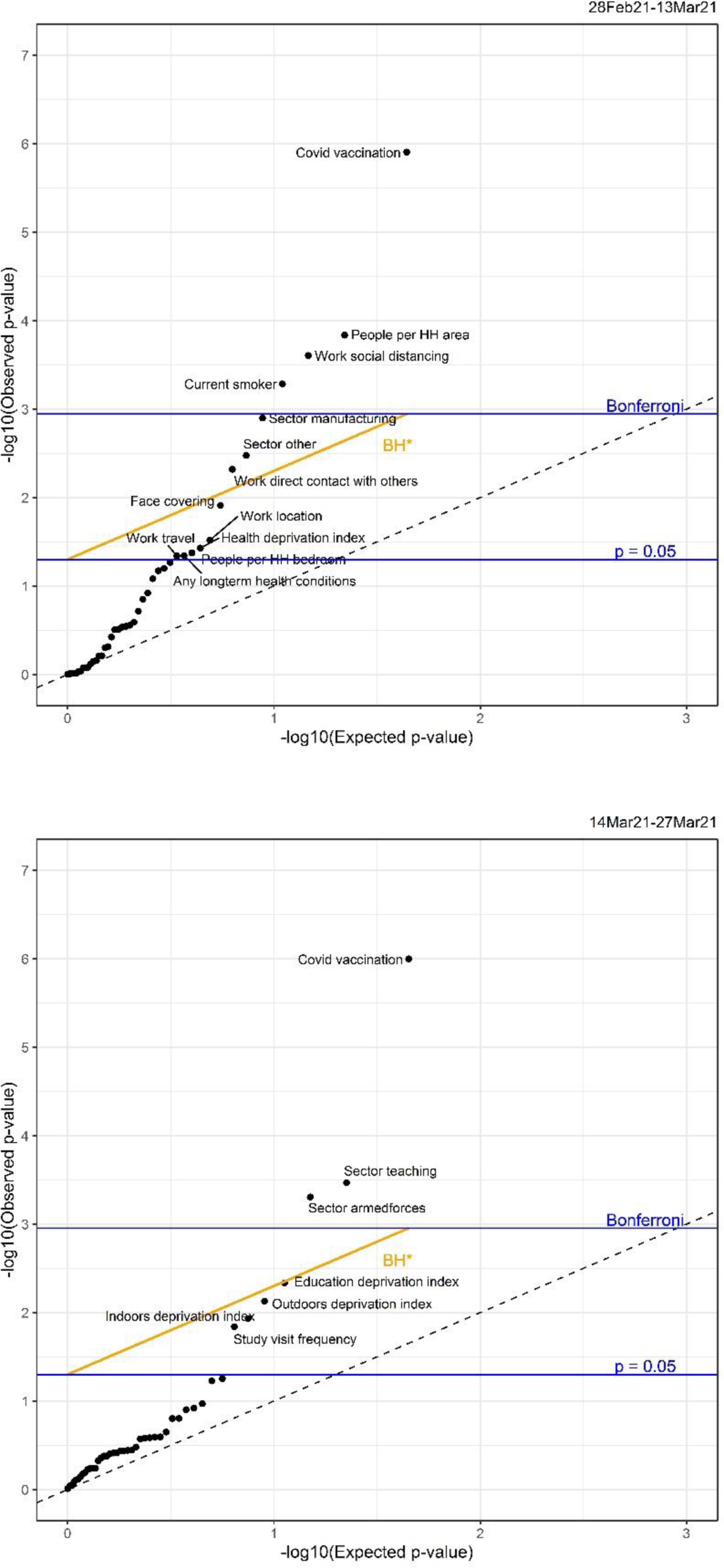
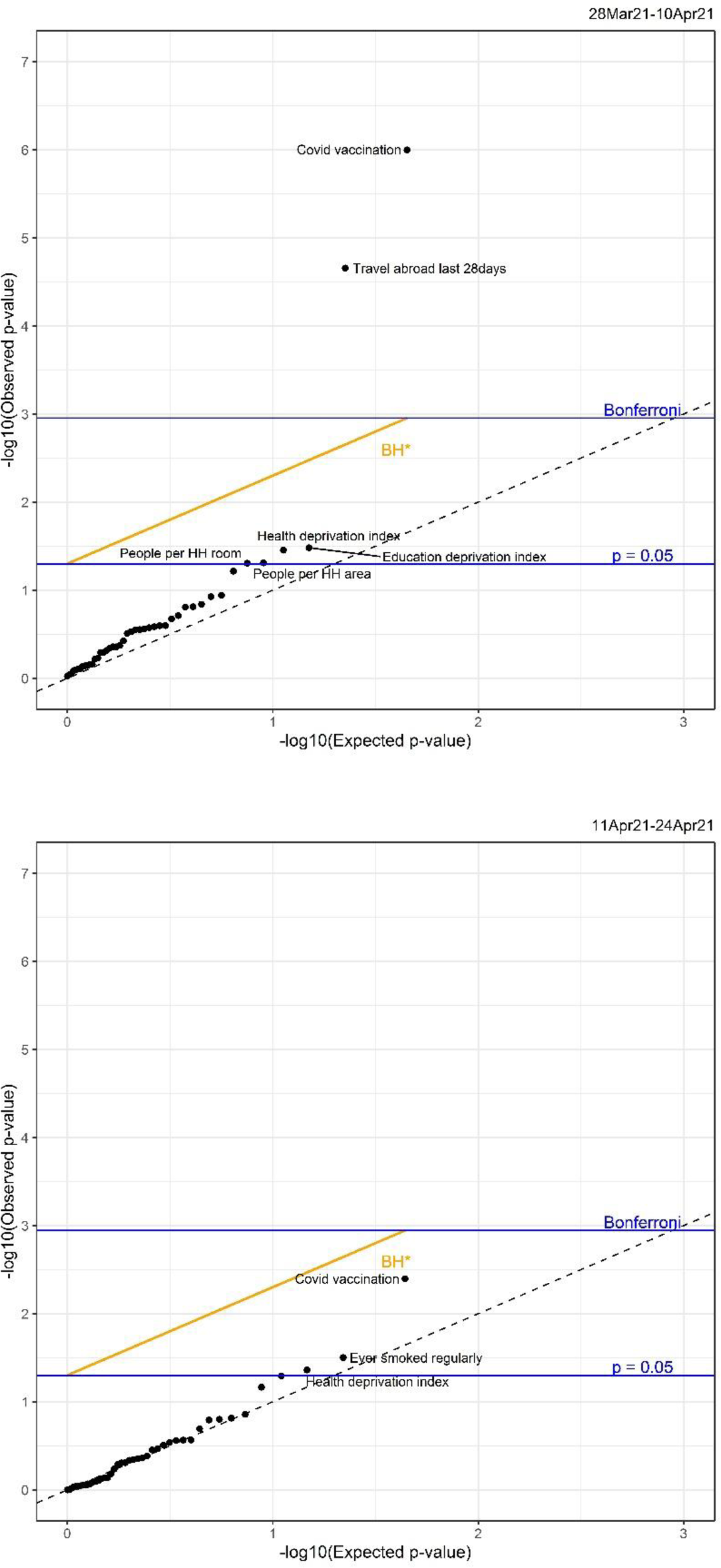
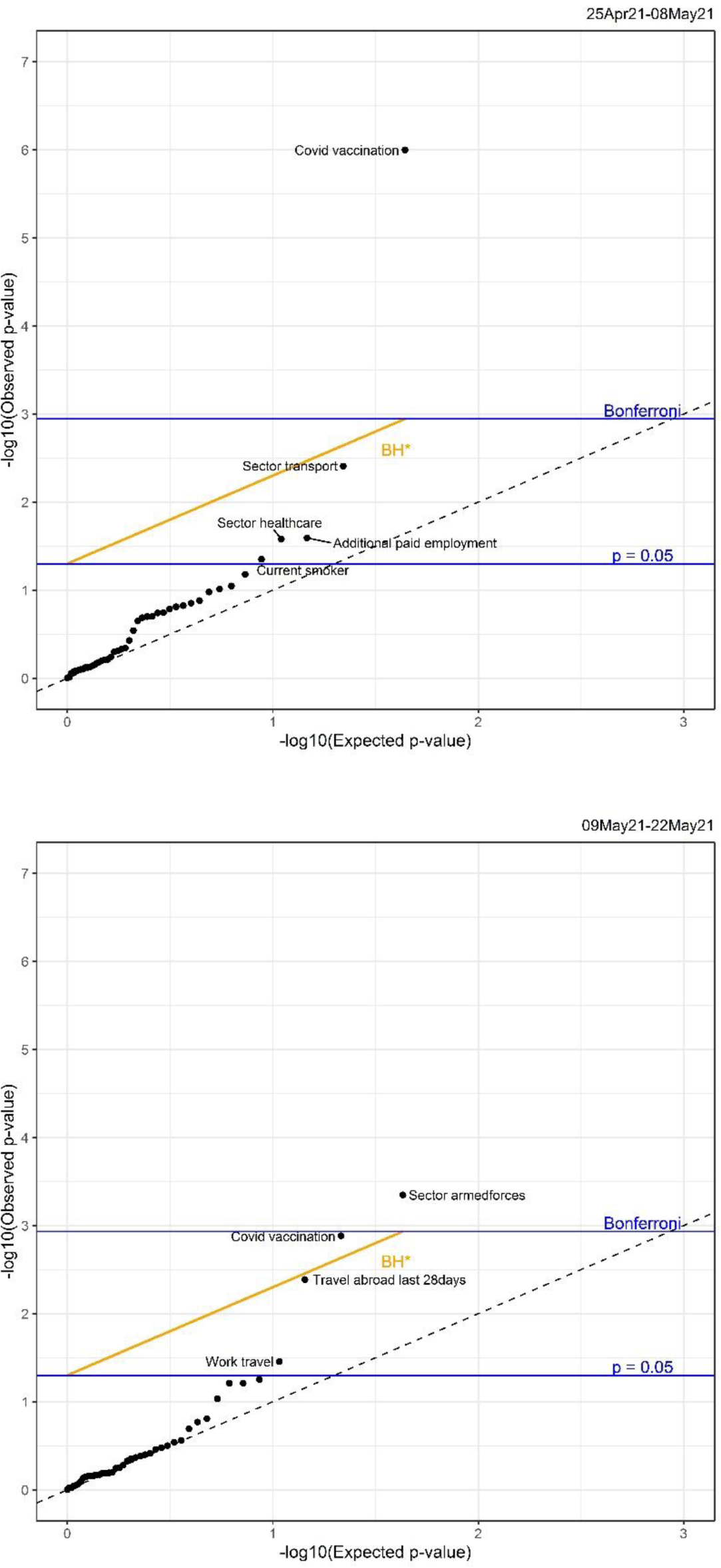
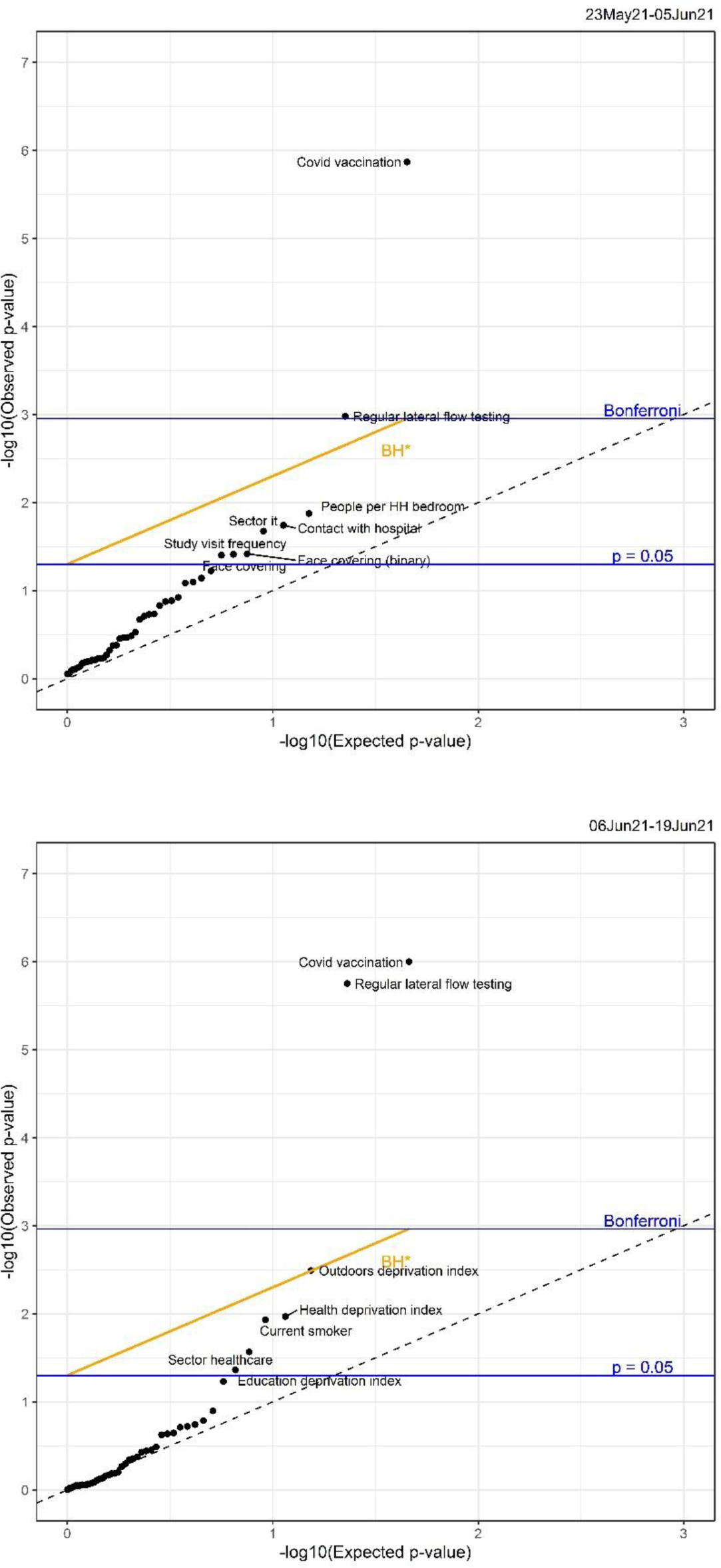
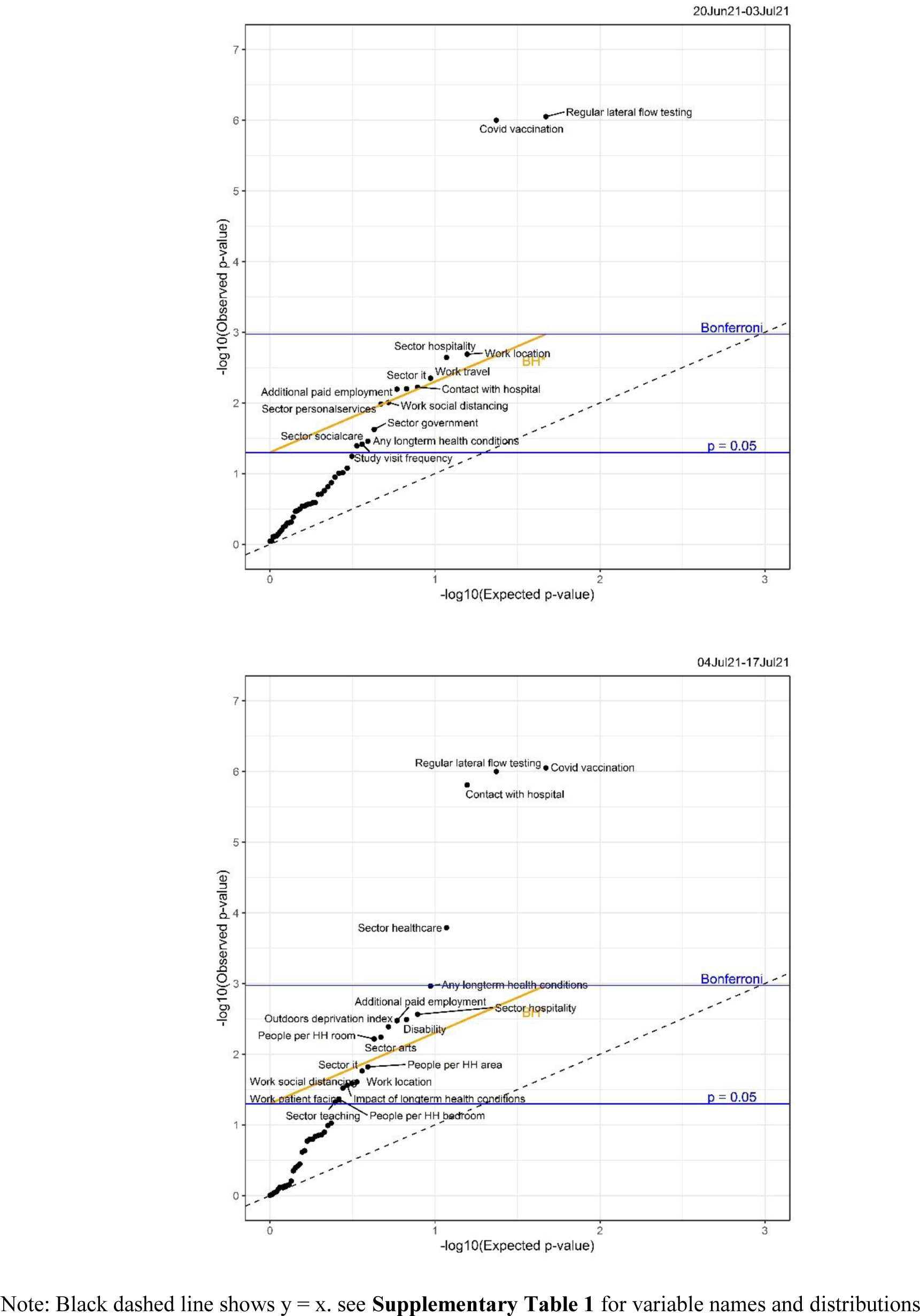
Global hetergeneity p-values per factor from the screening process over all 26 fortnights

